# Modeling of COVID-19 Transmission Dynamics on US Population: Inter-transfer Infection in Age Groups, Mutant Variants and Vaccination Strategies

**DOI:** 10.1101/2021.09.25.21264118

**Authors:** Jyotirmoy Roy, Samuel Heath, Doraiswami Ramkrishna, Shiyan Wang

**Affiliations:** Department of Chemical Engineering, Indian Institute of Technology, Powai, Mumbai 400076; Charles D. Davidson School of Chemical Engineering, Purdue University, West Lafayette, IN 47907

## Abstract

The in-depth understanding of the dynamics of COVID-19 transmission among different age groups is of great interest for governments and health authorities so that strategies can be devised to reduce the pandemic’s detrimental effects. We developed the SIRDV-Virulence epidemiological model based on a population balance equation to study the effect of mutants of the virus and the effect of vaccination strategies on mitigating the transmission among the population in the United States. Based on the available data from the Centers for Disease Control and Prevention (CDC), we obtain the key parameters governing the dynamic evolution of the spread of the COVID-19 pandemic. In the context studied, the results show that a large fraction of infected cases comes from the adult and children populations in the presence of a mutant variant of COVID-19 with high infection rates. We further investigate the optimum vaccine distribution strategy among different age groups. Given the current situation in the United States, the results show that prioritizing children and adult vaccinations over that of seniors can contain the spread of the active cases, thereby preventing the healthcare system from being overwhelmed and minimizing subsequent deaths. The model suggests that the only option to curb the effects of this pandemic is to reduce the population of unvaccinated individuals. A higher fraction of ‘Anti/Non-vaxxers’ can lead to the resurgence of the pandemic.

**Author summary:** The changing dynamics of the COVID-19 pandemic are primarily due to the mutations of the SARS-CoV-2 virus. It is often seen that these mutants not only have a higher infection rate but also evade the presently administered vaccines. To consider the fact that different age population groups are affected to varied extent by these mutants, we build a mathematical model to account for the inter-transfer infection among age groups, which can predict the overall COVID-19 transmission in the United States. The parameter quantification of our mathematical model is based on the public data for infected cases, deaths and vaccinated from the Centers for Disease Control and Prevention (CDC). Additionally, our study shows that the vaccine distribution strategies should be developed with a priority given to the most infected age groups in order to curb the total infected and death cases. We also show how the ‘Anti/Non-vaxxers’ can be a potential reason for resurgence of the pandemic. These results are of immediate practical application in determining future vaccine distribution regarding to the pandemic and ensuring the health care system is ready to deal with the worst-case scenario with a very high infection rate.

## Introduction

The coronavirus disease of 2019 (COVID-19) pandemic has been ravaging the entire world ever since its inception in Wuhan, China back in December 2019 [1]. The most challenging part of analyzing and predicting the COVID-19 pandemic in terms of infection and mortality numbers is its ever-changing dynamics due to the mutations of the virus [2]. For COVID-19, its mutations often cause an increase in the transmission rate of the virus [3] due to changes in its spike protein [2]. These mutations strengthen the interaction between COVID-19 virus and its receptor ACE2 [4]. This in turn also affects the vaccine efficacy [5] and thus creates a possibility of resurgence of the pandemic [6], which is revealed from the changing trends of the active infected cases and deaths in the US in recent times [7]. Since their peak in early January 2021, the COVID-19 cases and deaths had markedly declined, due in part to the increased vaccination coverage [8]. However, during June 19–July 23, 2021, COVID-19 cases increased approximately 30% nationally, followed by increases in hospitalizations and deaths [8], driven by the highly transmissible B.1.617.2 (Delta) variant, a variant of the severe acute respiratory syndrome coronavirus 2 (SARS-CoV-2).

The Delta variant is more than two times as transmissible as the original strains circulating at the start of the pandemic [9] and is causing large, rapid increases in infections, which puts pressure on the local and regional health care systems to provide medical care [9]. Strains on critical care capacity can increase COVID-19 mortality [10, 11] while decreasing the availability and use of health care resources for non–COVID-19 related medical care [12, 13]. In this scenario, it is important to predict the timing of peaks for active infections regarding worst-case scenarios, so that medical personnel are prepared and the Government can introduce appropriate and informed policy decisions.

One of the primary concerns is to identify the age groups which will be affected the most by delta strain of SARS-CoV-2 [14]. Age-related differences in responsiveness and tolerance become obvious and lead to worse clinical outcomes in elderly individuals [15]. Previous studies have mentioned that older COVID-19 patients are at an increased risk of death [16–19]. Accordingly, optimum vaccine strategies need to be developed, prioritizing the most affected age groups to reduce the peak of infected cases.

Despite widespread availability, vaccine uptake in the US has slowed nationally with wide variation in coverage by state (range = 33.9%–67.2%) and by county (range = 8.8%–89.0%) [20]. Unvaccinated persons, as well as persons with certain immunocompromised conditions [21], remain at substantial risk for the infection, severe illness, and death, especially in areas where the level of SARS-CoV-2 community transmission is high [22]. Therefore, the proportion of people who are not willing to get vaccinated is significantly affecting the future dynamics of the COVID-19 pandemic. Although a study found a 67% acceptance of a COVID-19 vaccine, there were noticeable demographic and geographical disparities in vaccine acceptance [23]. With increasing anti-vaxxer (people who are opposed to vaccination) rhetoric towards the COVID-19 vaccines that have been developed, the resultant dynamic evolution of COVID-19 transmission becomes more complicated. While the government has focused on reducing the spread of misinformation and addressing the underlying concerns of the anti-vaxxer community [24], it would be important to provide a quantitative understanding of the effect of anti-vaxxers on the dynamics of COVID-19 transmissibility among the community.

The COVID-19 disease has affected different age groups to different extents. Studies by Chikina et al. have developed SIR-like epidemic models integrating known age-contact patterns for the United States to model the effect of age-targeted mitigation strategies for a COVID-19-like epidemic [25]. It has been seen that strict age-targeted mitigation strategies have the potential to greatly reduce mortalities and ICU utilization. Another study by Yu et al. reaffirmed that COVID-19 epidemic processes have had distinctive dynamic patterns among age and gender group [26]. The epidemic among young adults led the epidemic process across the whole population, with a second peak occurring in people aged 20–39 years. Studies by Moghadas et al. have highlighted the importance of incorporating mutations and evolutionary adaptations in epidemic models [27]. They also demonstrated how the multiple-strain transmission model can be used to assess the effectiveness of mask-wearing in limiting the spread of COVID-19. Thus, in this scenario vaccination strategies become of primary importance. Vaccination markedly reduced adverse outcomes, with non-ICU hospitalizations, ICU hospitalizations, and deaths decreasing by 63.5% (95% CrI: 60.3% - 66.7%), 65.6% (95% CrI: 62.2% - 68.6%), and 69.3% (95% CrI: 65.5% - 73.1%) [28], respectively, across the same period. To date, there is an urgent need for studies focusing on the effects of different age groups on the COVID-19 transmission dynamics with mutual considerations of mutation and vaccination strategy. In this work, we model the interaction among the age groups as the infection brought by the virulence environment [29, 30]. We correlate factors of variants and vaccination efficacy by studying the effect of mutants on various age groups and devise vaccine distribution strategies.

In this study, we will apply the population balance model to derive the average equation, which is used to simulate the COVID-19 transmission that has occurred in the United States. We will focus on three aspects of COVID-19 transmission modeling, as illustrated in figure 1(a). Firstly, we will evaluate how an increased transmission rate of delta variant and the increased vaccine inefficacy due to the mutation can change the dynamics of the pandemics in the US. We will identify the age groups which are supposed to be affected the most by this new strain. Secondly, we will attempt to design an optimum vaccine distribution strategy prioritizing the most affected age groups so as to bring down the active infections and mortality. Thirdly, we will account for the effect of anti-vaxxers to determine what proportion of population needs to be vaccinated to prevent the resurgence of the pandemic.

**Fig 1.**
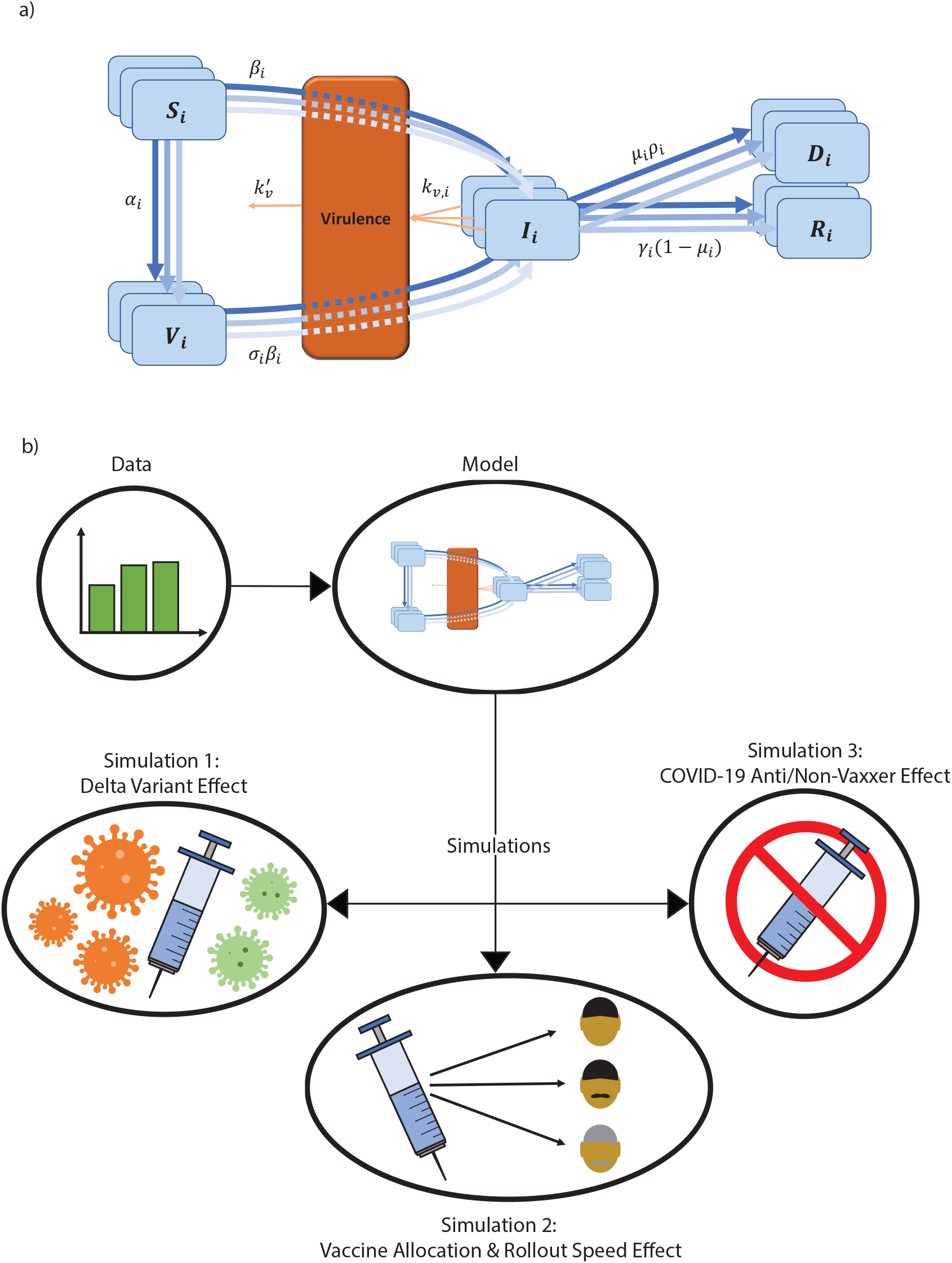
a) An SIRDV-Virulence compartmental model to predict the transmission of COVID-19 in the United States, in which the members of three distinct age groups move to compartments (blue) at the rates given adjacent to the inter-compartmental arrows (blue). We consider the infection inter-transfer frequency among three different age groups (children, adults, and seniors). Infected members of each age group contribute to a single virulence parameter (orange), which can infect both susceptible, (*S*_*i*_), and vaccinated, (*V*_*i*_), individuals. b) Data is fed to the compartmental model to fit its parameters. The fitted model is used to run simulations to predict future scenarios: 1) the effect of the delta variant, 2) the effect of changing the vaccination allocation and the vaccination roll-out speed, and 3) the effect of changing the proportion of COVID-19 Anti/Non-Vaxxers.

## Materials and methods

Our model considers the interactions among different age groups using population balance equations [31, 32]. The subsequent average equations are fitted under the collected COVID-19 transmission data. In the following sections, we will start with the introduction of the model for the infection transfer among three different age groups (i.e. children, adult, senior); then derive the average equations using the population balance model, which can be converted to dimensionless equations with characteristic quantities (dimensionless numbers); we quantify the dynamic of these dimensionless numbers using CDC released infection, death, and vaccination data.

### Compartment model

In figure 1(b), our SIRDV-Virulence model consists of 6 compartments: Susceptible (*S*_*i*_), Infected (*I*_*i*_), Recovered (*R*_*i*_), Dead (*D*_*i*_), Vaccinated (*V*_*i*_), and virulence (𝒱), where *i* denotes different age groups (1: children, 2: adult, and 3: senior). To illustrate the interactions among different compartments and age groups, we start by introducing the average infection transfer to characterize the interactions among the different age groups. The dynamics equations can be formulated by the population balance equation.

### Average infection inter-transfer frequency between any two age groups

Here, we identify the age-specific infection transfer frequency as *q*(*τ, τ*′; *t*)*dt* in figure 1(b), which is the probability that two age groups *τ* and *τ* ′ who are in each other’s neighborhood will meet with the other in closed proximity during the time interval *t* to *t* + *dt*. For instance, we choose *τ* to be in the children range denoted as *g* = *S*_1_ (*S*: Susceptible) and *τ*′ to be any other age range (e.g. adult, elderly) identified as *g*′ ∈ {*I*_2_, *I*_3_} or self *g*′ = *I*_1_ (*I*: Infected). The average meeting frequency between individuals in *g* and *g*′ is identified as:

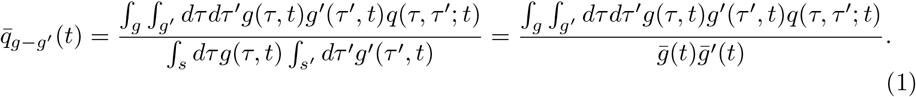

### Population balance model of infection dynamics

A model considers the transfer of infection among the children, adult and senior age groups. Note that this model can be extended to any number of groups. First we must add two internal coordinates where *v* denotes the viral population in the individual and *a* represents the antibody population. Thus, we let *ϕ*(*v, a, t*)*dvda* be the number of individuals with a viral infection in the range *v* to *v* + *dv* and an antibody in the range of *a* to *a* + *da*. The viral infection in each individual changes at the averaged rate 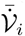 in population *i*, where 𝒱(*t*) = ∫_*v*_ *dv* ∫_*a*_ *daϕ*(*v, a, t*) is denoted as virulence. Note that, in this work, we focus on the effect of virulence environment among population interactions and neglect the factor of viruses resident on the solid surfaces and droplets [33]. Therefore, this definition of virulence 𝒱 excludes viruses resident on the solid surfaces in the environment, which can also cause spreading to the extent that people come into contact surfaces.

We note the total population density of infected individuals *I*_*i*_,

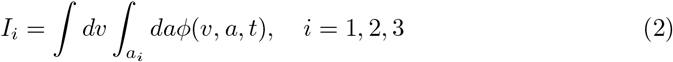

where *a*_*i*_ indicates the age range of group *i*; we assume that a freshly infected individual has *v* = 0 and *a* = 0. Then, 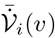 is the viral infection change rate in the individual and 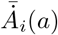 is the antibody concentration change rate in the individual. An infected individual with viral intensity *v* may die with the frequency *k*_*d*_(*v, a*) or recover with frequency *k*_*r*_(*v, a*). The population balance equation for the diseased population is given by

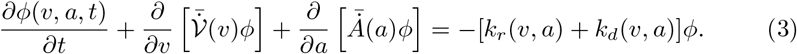

We denote the existence of probability densities *p*_*i*_(*v*) in age group *i* for developing an infection on contact with an infected individual of viral density *v*, 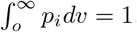. The boundary condition for equation (3) is

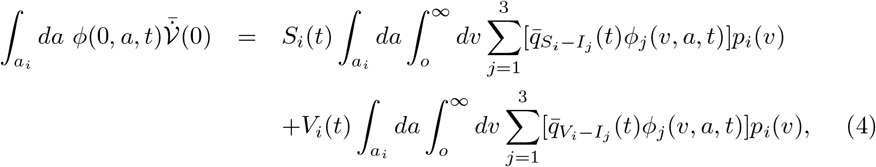

where *S*_*i*_ is the susceptible population, *V*_*i*_ is the vaccinated population, and 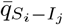 is the meeting frequency between susceptible *S*_*i*_ and infected *I*_*j*_.

### Average Models

We further observe that the total infection equation obtained by integrating equation (3) with respect to *v* from zero to infinity and *a* within the age range (*a*_*i*_),

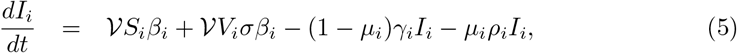

where we applied the boundary condition equation (4) and 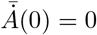; the transmission rate *β*_*i*_ and vaccine efficacy *σ* are thus defined as

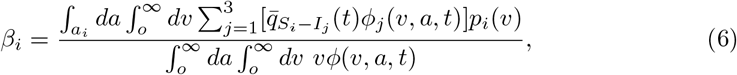

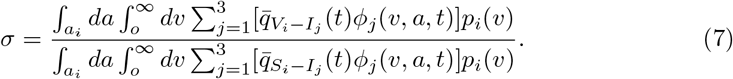

The rate constant *β* characterizes the extent of the inter-transmission among different age groups, and *σ* measures the ratio of transmission rate between vaccinated and susceptible populations. The death and recovery rates are expressed as

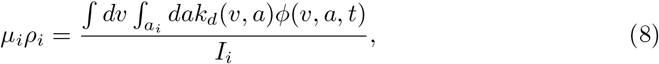

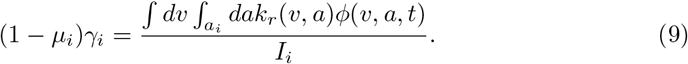

Next, we multiply equation (3) by *v* and integrate with respect to *v* and *a* from 0 to infinity:

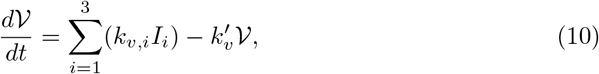

where

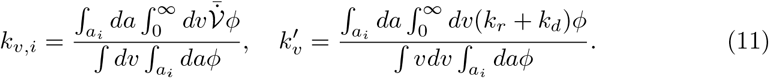

With dynamics of virulence 𝒱 and infection *I*_*i*_, it would be straightforward to derive the equations for the rest of compartment, which has been displayed in supplementary material.

### Dynamics Models

To understand both dynamics of viral and infected & vaccinated populations, the modeling of the system in a considered geometric domain was abstracted as its dimensionless form:

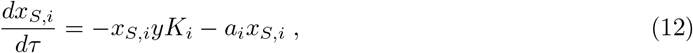

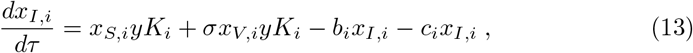

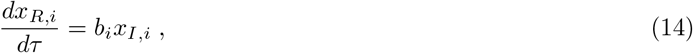

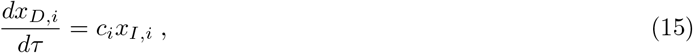

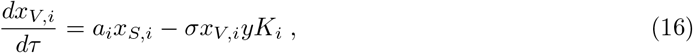

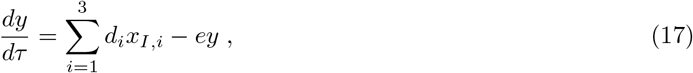

where a list of dimensionless parameters is defined in Table 1. The susceptible fraction for age group *i* is represented by *x*_*S,i*_; in figure 1(b) and equation (12), people from this compartment either move into the infected compartment (represented by *x*_*I,i*_), or into the vaccinated compartment (represented by *x*_*V,i*_). The infection transition rate from the susceptible compartment is characterized by the dimensionless transmissibility of the virus (*K*_*i*_) and the viral load (*y*). On the other hand, the dimensionless vaccination rate (*a*_*i*_) determines the rate of transition from susceptible to vaccinated compartment. For the dynamics of the infected compartment (Eq (13)), the total rates are determined by the rate of infection of susceptible and vaccinated populations, as well as that of recovery & death. Since none of the COVID-19 vaccines currently being administered have 100% efficacy [34], there is a probability of vaccinated people getting infected, which is governed by vaccine inefficacy, *σ*. From the infected compartment, a person can either move into the recovered compartment or the dead compartment, which are governed by dimensionless recovery rate (*b*_*i*_) and dimensionless mortality rate (*c*_*i*_), respectively (see equations (14)-(15)). In equation 16, the population in the vaccinated compartment is governed by the vaccination rate and the vaccine efficacy. Lastly, the dynamics of virulence are governed by the dimensionless number *d*_*i*_, which is contributed by the infected compartment. There is also a removal term from the viral load compartment dictated by dimensionless number *e*, which is related to the lifespan of the virus.

**Table 1.**
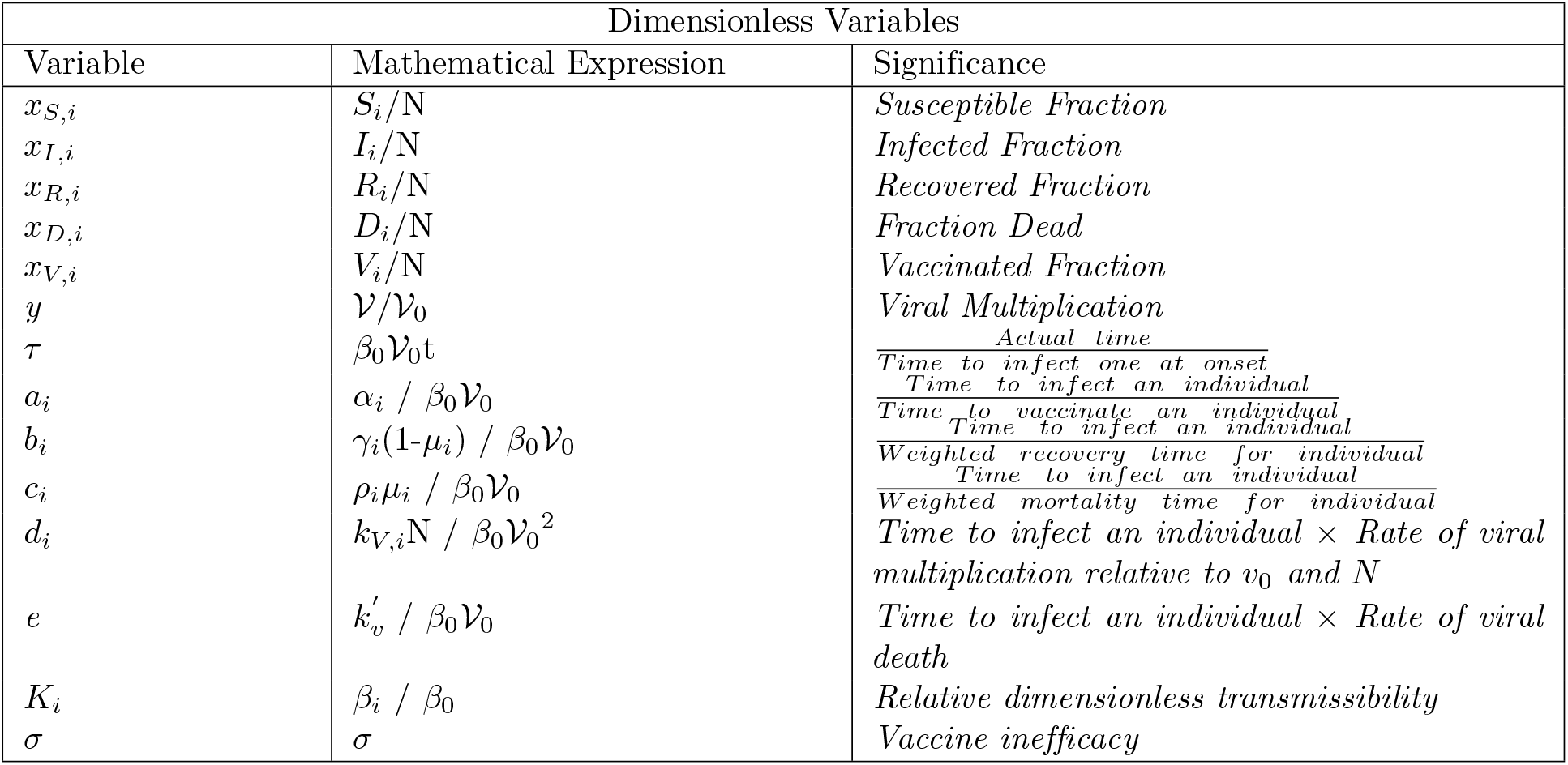
Definition of variable for the dimensionless SIRDV-Virulence model

### Data Processing

We considered three different age groups for analysis: children (12-17 years), adults (18-64 years), and seniors (65 years and older). These age group divisions mirrored those of the CDC’s vaccination data set [35]. Because the infected cases and death data sets used for this study had smaller age group divisions than those of the vaccination data set, the age group data of each data set were combined to form uniform age groups across all three data sets (figs. S1-3 in supplementary material). To analyze the COVID-19 dynamic evolution, we quantified dimensionless quantities based on data of infection, death and vaccination, which was normalized by the total population. For the analysis, the total population of the United States was assumed to be 332.5 million [36], based on the current US population at the time of data collection. The United States Census 2019 age distribution [36] was used to estimate the population of age groups in the United States. Because each data set used in this study had a unique format, the data sorting and processing for each set were different, as described in the supplementary material.

### Simulation Scheme

Before fitting the dynamic SIRDV-Virulence equations (12)-(17) to the case, death, and vaccination data, it was necessary to estimate the initial value for each compartment. The method for estimating the initial value of each compartment has been outlined in supplementary materials, which was repeated for each of the three age groups.

Note that data were not available for the susceptible population. Since the Susceptible compartment is the final compartment under consideration, a population balance was used to calculate this value:

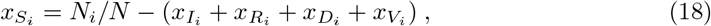

where *N*_*i*_ is the population of age group *i* and *N* is the total population (ages 12 and older). For simplicity, it is assumed that the population of each age group is constant during the time period for fitting the model and completing the future predictions. The youngest age being considered is 12 years old, validating the assumption that births will not affect the change in population sizes, and deaths from causes other than COVID-19 are considered negligible. Further, monitoring the changes in population as a result of movement of international migrants into and out of the United States population (among other population dynamics) is considered negligible and outside the scope of this work.

We non-dimensionalize the raw data so that it would be consistent with the dimensionless equations used in the model. When the SIRDV-Virulence model was used for fitting individual age group dynamics, the data were normalized with the total population of each respective age group, i.e, children, adults or seniors. When simultaneously fitting the dynamics of all age groups, the data were normalized using the total United States population of the ages of 12 years and older, i.e, a summation of the population of children, adults and seniors.

## Results

### Parametric Study for different infection period

Since the vaccine dynamics and effect of mutant variants of the virus varied from January to July of 2021, four different time periods were considered for fitting and the 16 parameters (*K*_*i*_, *a*_*i*_, *b*_*i*_, *c*_*i*_, *d*_*i*_, *e* with *i* = 1 *− children*, 2 *− adults*, 3 *− seniors*) were obtained for each of these time periods in figure 2(a). Three data sets were used for model fitting: case data (2b), death data (2c), and vaccination data (fig. S4 in supplementary material). The fitted model against the data sets for each time period can be seen in Figures S6-11 in the supplementary material. The first time period considered was from January 9, 2021 to March 6, 2021. During this period, the most transmissible strain of COVID-19 present was the alpha variant [37]. The children were not vaccinated during this time period and only the seniors and adults were vaccinated. The next period considered was from March 6, 2021 to May 8, 2021. In this time period, it was assumed that the most transmissible strain of COVID-19 present was the delta strain. This assumption is logical as the CDC website [38] also reports the introduction of delta variant in the US in the early March of 2021. During this period the vaccination status remained the same as the previous time period, i.e, the adults and seniors were vaccinated and the children were not. The third time period was from May 8, 2021 to June 12, 2021. Within this phase, all three age groups (children, adults and seniors) were vaccinated. The alpha and delta variants were considered the most dominant strains, and the delta variant was beginning to contribute to a significant proportion of recorded cases [39]. There was a moderate decrease in new cases and deaths during this period as seen in figures 2(b,c). The last period was from June 12, 2021 to July 31, 2021. During this period, the proportion of cases due to the delta variant increases as indicated by an increase in the number of cases. During this period, the vaccination rates were faster for children as compared to adults and seniors (shown in 2(e)). Thus, 4 sets of 16 parameters were obtained for these four time periods.

**Fig 2.**
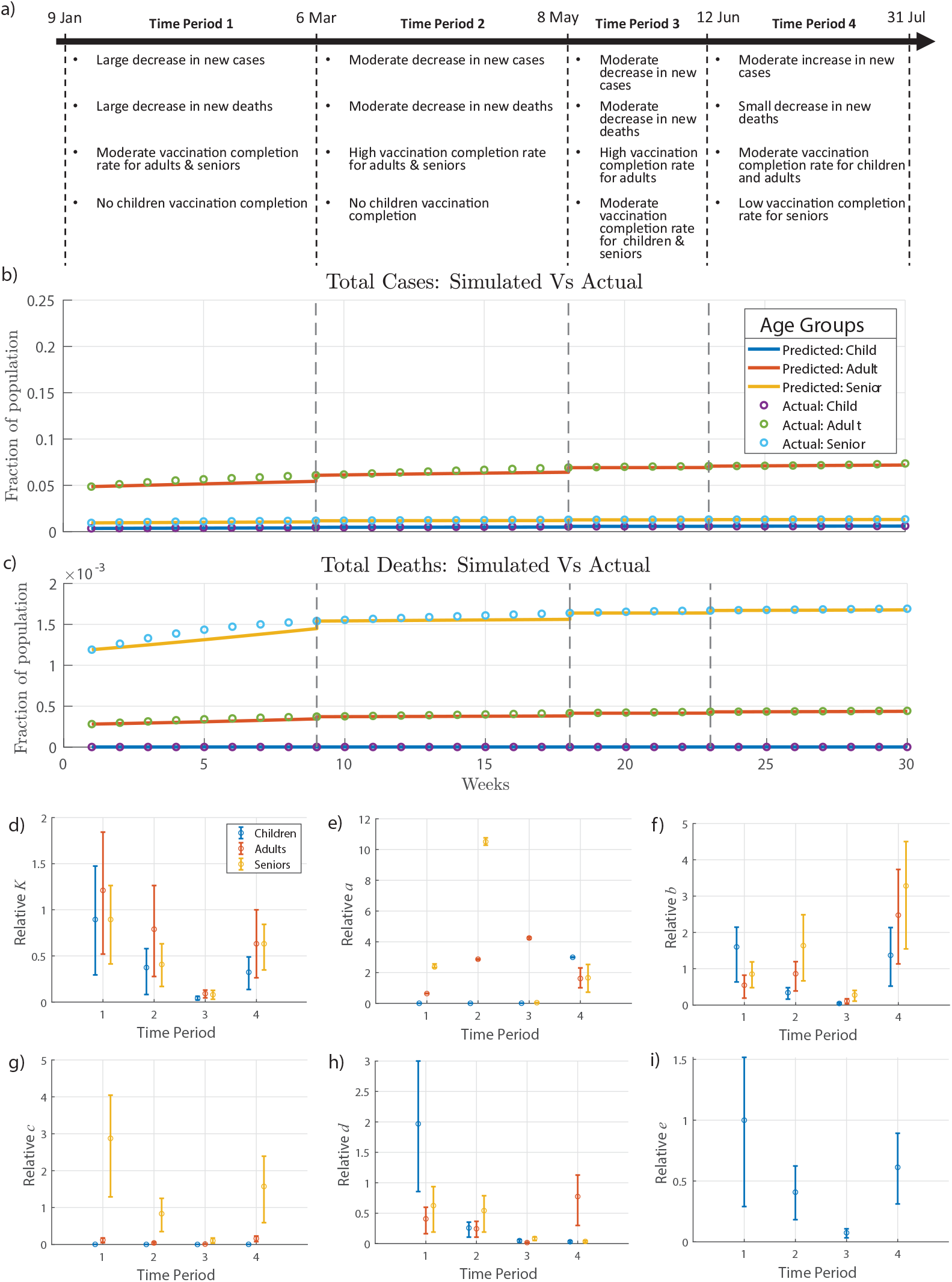
SIRDV-Virulence Model Fitting. (a) COVID-19 data are divided over four time periods based on the characteristics of the data sets from the CDC in the United States. The fitted model is compared to the data during each time period for (b) cumulative weekly cases, (c) cumulative weekly deaths, and cumulative weekly vaccinations (shown in the supplemental material). Quantified dimensionless parameters are shown for each age group (i=1-children,2-adults,3-seniors) during each of the four time periods: (d) viral transmission *K*_*i*_, (e) vaccination rate *a*_*i*_, (f) recovery rate *b*_*i*_, (g) death rate *c*_*i*_, (h) viral load *d*_*i*_, and (i) life span of virus *e*. In figures (d-i), the mean is the central point, and the error bars represent the 25^*th*^ and 75^*th*^ percentile values; relative parameters are displayed, where each data point is relative to the average of the respective parameter values of the three age groups for the first time period.

The quantified dimensionless numbers have properly reflected the dynamics of COVID-19 transmissibility during these four periods. In figure 2(d), the value of relative transmission rate (*K*_*i*_) was showing a decreasing trend for all three age groups during periods 1-3. However, as the delta variant has progressed to a large fraction of population, there is a sudden increase in the values of *K* for all age groups in the period 4 (fig 2d).

The dynamics of vaccination rate *a*_*i*_ correctly captures the current vaccination strategy in the United States. During the first two time periods, *a*_*i*_ was higher for the senior age group (*i* = 3), which is consistent with prioritizing senior vaccination. In the last two periods (figure 2e), the children and adult age group, on average, have a higher vaccination rate than the senior age group.

The recovery rates for all the three age groups shows an increasing trend among all three age group as time progresses. This is indicated in figure 2f. The main reason behind this, with a higher fraction of the population getting vaccinated with time, the immunity is expected to increase. However, there is an increase in the mortality rate of the senior age group from the third to the fourth time period as indicated by a high value of *c* in figure 2g, which can primarily be due to the delta variant effect. Relative to the senior age group, the changes in mortality rates of children and adults are negligible, even during the last period when the delta variant effect is visible in the population. The adult age group contributes the most to the viral load during the last period as indicated by a high value of *d*_*i*_ in figure 2h. The dimensionless number *e* shows a decreasing trend followed by a sharp increase in the last time period as seen in figure 2i. This indicates the fact that the time to infect an individual times the viral death rate increases when the delta variant effect is visible.

In the following sections, we will discuss the future predictions considering three important factors: (1) effect of the delta variant, (2) vaccine optimization, and (3) effect of Anti/Non-Vaxxers.

### Effect of mutation on transmissibility

The mutation of the virus has been largely responsible for the increase of transmissibility due to increased infection rate and reduced vaccine efficacy [40]. Recently, the delta variant is responsible for almost all recorded COVID-19 cases [7], in accordance with the elevation of infection (*K*_*i*_) in the fourth fitted time period. Therefore, it is important to study the effects of changes in terms of *K*_*i*_ and vaccine efficacy *σ* on the future active number of cases and number of deaths. First, we study the variation in *K*_*i*_ while keeping vaccine inefficacy, *σ*, constant. The simulated values of infection rate, *K*_*i*_, were taken to be 1, 1.2, 1.5 and 2 times the *K*_*i*_ values from the fourth period (June 12, 2021 to July 31, 2021). It was assumed that the same amount of increase takes place for the *K*_*i*_ belonging to different age groups. For all the age groups in figures 3(c-e), the number of active cases increases with the elevating *K*_*i*_. For instance, if there is a two fold increase in *K*_*i*_, the number of active cases at the peak of pandemic increase by around 1.5 to 2 times for all age groups (see *K*_*new*_*/K* = 2 in figures 3(c-e)). In addition, a larger infection rate (*K*_*i*_) would delay the moment when the infection reaches the peak values. We estimate that the peak for adult and senior age group is expected sometime around October 2021, while for children it is expected around December 2021, which reveals various infection dynamics for different age groups. Similarly, the total number of deaths increases with an increase in the value of *K*_*i*_ for all age groups (figures 3(f-h)). The effect is more pronounced in children and adult category as compared to the senior category, which is due to larger numbers of unvaccinated individuals in both children and adult populations comparing to seniors. For a two fold increase in *K*_*i*_, the total number of deaths goes up by approximately 8% in children and 10% in adults, whereas in senior the increase is around 2%.

**Fig 3.**
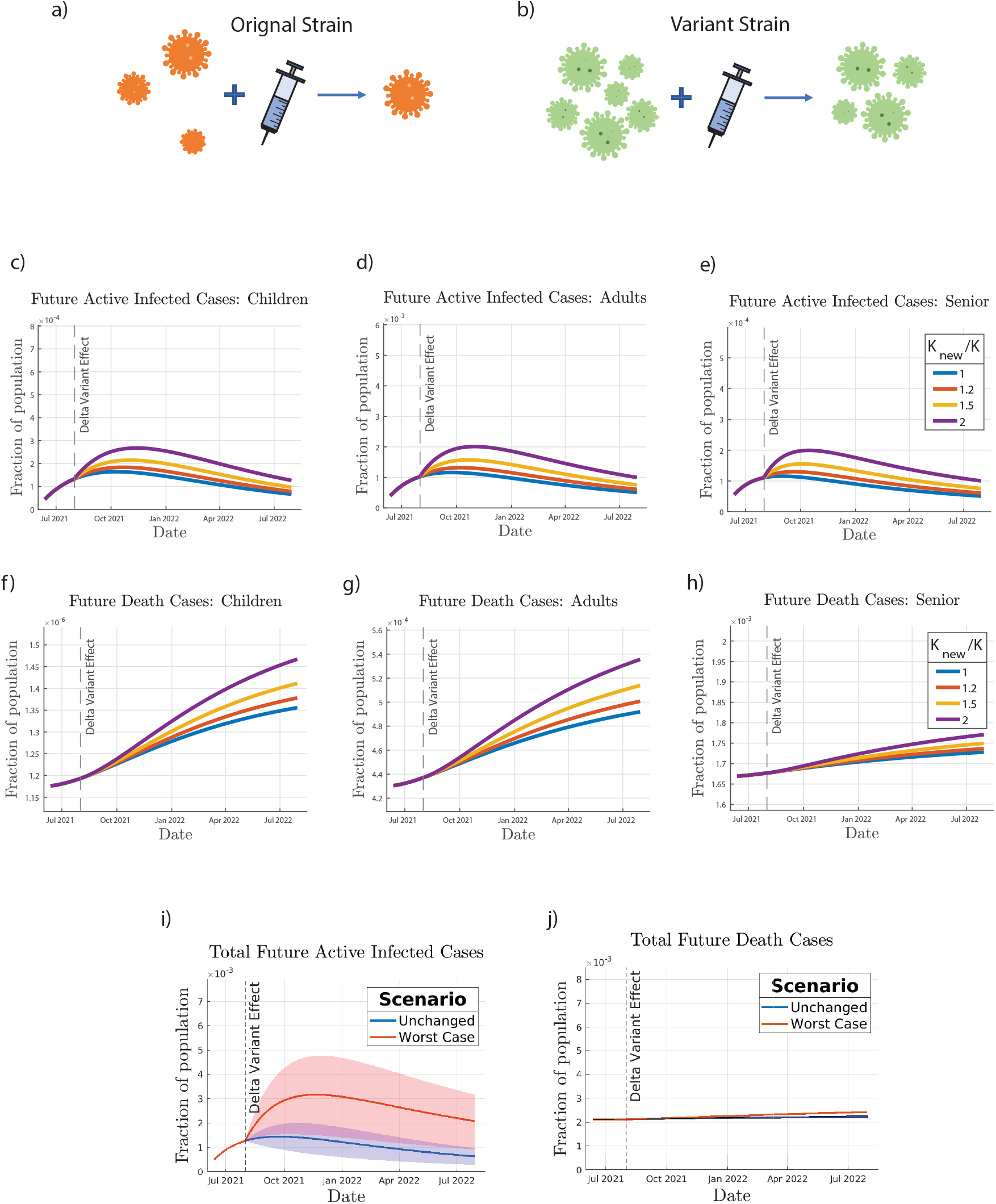
Effect of mutation on COVID-19 transmission. The mutant variant is modeled by assuming an increase in infection rate *K*, and an increase in vaccine inefficacy *σ* by comparing (a) original and (b) variant strain, shown by a higher concentration of virus particles and a higher proportion of particles after vaccination. Future predictions of infected cases and deaths are simulated for (c,f) children, (d,g) adults, and (e,h) seniors at increasing relative infection rate (relative to *K* from the fourth fitted time period) and a constant *σ* of 0.05. The worst case scenario (relative *K* of 2 and *σ* of 0.2) is simulated, resulting in the predictions of (i) total future active infected cases and (j) total future deaths, where the shaded regions show the error introduced using the 25^*th*^ and 75^*th*^ percentiles of the relative *K* values.

A comparison was made between two extreme scenarios in figure 3(i,j), where we compare no change in dimensionless numbers (from the fourth fitted time period) versus the worst case scenario. In the worst case scenario, not only is the relative dimensionless infection rate doubled, but the vaccine inefficacy is increased from 0.05 to 0.2. While the total number of deaths is not significantly affected in the worst case scenario, the active infected cases increases by almost 2.5 times with the peak occurring sometime around December 2021 for all the age groups combined.

Further simulation scenarios can be found in the supplemental material: variation of *K* with *σ* = 0.20 (figs. S12-13), variation of *σ* with *K*_*new*_*/K* = 1 (figs. S14-15), and variation of *σ* with *K*_*new*_*/K* = 2 (figs. S16-17).

### Vaccination optimization strategy

Optimization of vaccine distribution strategies among different age groups remains critical [41]. Specifically, it would be interesting to study the effect of varying the vaccination rates and vaccination prioritization among each of the three age groups. Our study modeled the resulting completed vaccinations, cumulative cases, and cumulative deaths over a future time period as the vaccination parameters were varied.

To determine the practical range for the dimensionless vaccination parameter *a*_*i*_ of each age group, we need to consider two constraints. Firstly, the maximum completed vaccination rate for future simulations for a single age group was set as the maximum weekly completed vaccination rate achieved during the fitted time period for the respective age group (fig. S5 in supplementary material). Secondly, the maximum completed vaccination rate for future simulations for all age groups combined was then set as the maximum weekly completed vaccination rate during the fitted time period. In other words, the predicted simulations were completed under the assumption that future vaccination rates in the United States would not reach the rates that they had reached previously (considering both individual age groups and the entire population under study), given the majority of the senior and adult age groups had already been vaccinated by the initial date of the future simulation time period and peaks had already been reached for the completed vaccination rates in each age group.

In figure 4, a comparison was done for vaccination priority for age groups with the heat maps. In general, the minimum infected cases and deaths and the maximum fraction of vaccinated population occur at the highest values of *a*_*i*_ for these age groups, yet the dependence of vaccinate rate in each age group is different. For instance, the future total infected and deaths, as well as total vaccinated fraction are more dependent on *a*_2_ (adult age group) than on *a*_1_ (children age group) as shown in figures 4(a,b). This is mainly because the fraction of the adult population is much higher than that of children. For the comparison among children and senior age groups, it was seen that the total death and infection were more strongly dependent on *a*_1_ than on *a*_3_ (senior age group) as seen in figures 4(d,e). This is because a large fraction of seniors (∼ 81.8%) [42] had already been fully vaccinated for COVID-19. In comparison, only around 54.4% of the adult population was vaccinated by the end of July 2021, while the percentage of children fully vaccinated was even lower (∼ 34.4%) [42].

**Fig 4.**
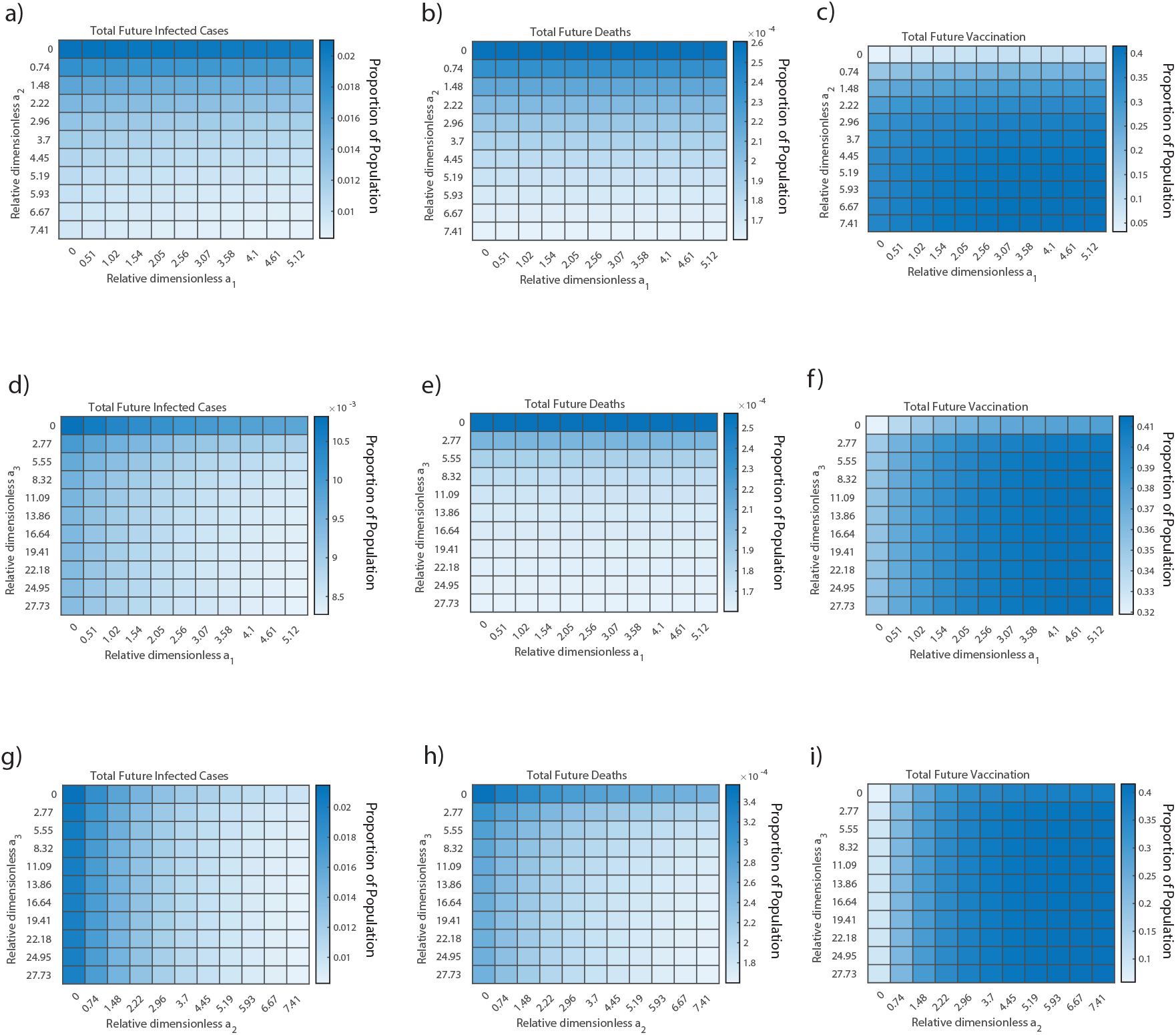
Vaccination Allocation Results. The effect of changing the vaccination allocation among each age group and vaccination roll-out speed is studied. (a,b,c) The senior vaccination parameter, *a*_3_, is held at its maximum practical value, while the children and adult vaccination values, *a*_1_ and *a*_2_, respectively, are varied from 0 to their maximum practical values to predict the proportion of the United States population that will (a) become infected and (b) die as a result of COVID-19 from July 2021 to July 2022. (c) The total number of completed vaccinations from July 2021 to July 2022 as a result of the same parameter changes are shown for reference. (d,e,f) The adult vaccination parameter is held at its maximum value while the children and senior vaccination parameters are varied. (g,h,i) The children vaccination parameter is held at its maximum value while the adult and senior vaccination parameters are varied.

Since a large fraction of children population is yet to be vaccinated, a higher priority needs to be given to the children age group over the senior age group for the future vaccine distribution, which is consistent with the current ongoing strategy in the United States [43]. A similar comparison among the senior and adult age group showed that total death and infected cases is more dependent on *a*_2_ than *a*_3_ as seen in figures 4(g,h). The reason is same as the previous case: a large fraction of adult category had yet to be vaccinated by the end of July 2021. So the observations from these heat maps led to the conclusion that, for the vaccine distribution strategy, a higher priority needs to be given to adult and children categories than senior category to minimize total death and infection cases as the majority of population in these two groups are more susceptible to the infection.

### Effect of Anti/Non-Vaxxer

The objective of the third simulation case was to determine the long-term effects of individuals unwilling and/or unable to receive the COVID-19 vaccination(s). Specifically, the effects of varying the proportion of COVID-19 ‘Anti/Non-Vaxxers’ in the susceptible population of each age group were observed to see how changes in the proportion of one age group could affect the number of deaths and cases of that same age group or other age groups.

To simulate this case, a modified compartmental model, PAIRDV-Virulence, was developed that divided the Susceptible compartment into two new compartments: the COVID-19 ‘Anti/Non-Vaxxer’ compartment and the COVID-19 ‘Pro Vaxxer’ compartment, as shown in figure 5(a). The relationship between the Susceptible compartment of the SIRDV-Virulence model and the COVID-19 ‘Anti/Non-Vaxxer’ and COVID-19 ‘Pro Vaxxer’ compartments of the PAIRDV-Virulence is 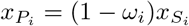 and 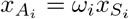, where 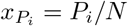 represents the number of individuals of age group *i* in the COVID-19 ‘Pro Vaxxer’ compartment, and 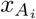 represents the number of individuals of age group *i* in the COVID-19 ‘Anti/Non-Vaxxer’ compartment. At the beginning of the time period of future simulations, 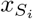 is the sum of 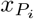 and 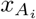, and *ω*_*i*_ is the proportion of COVID-19 ‘Anti Vaxxers’ in the susceptible population of age group *i*.

**Fig 5.**
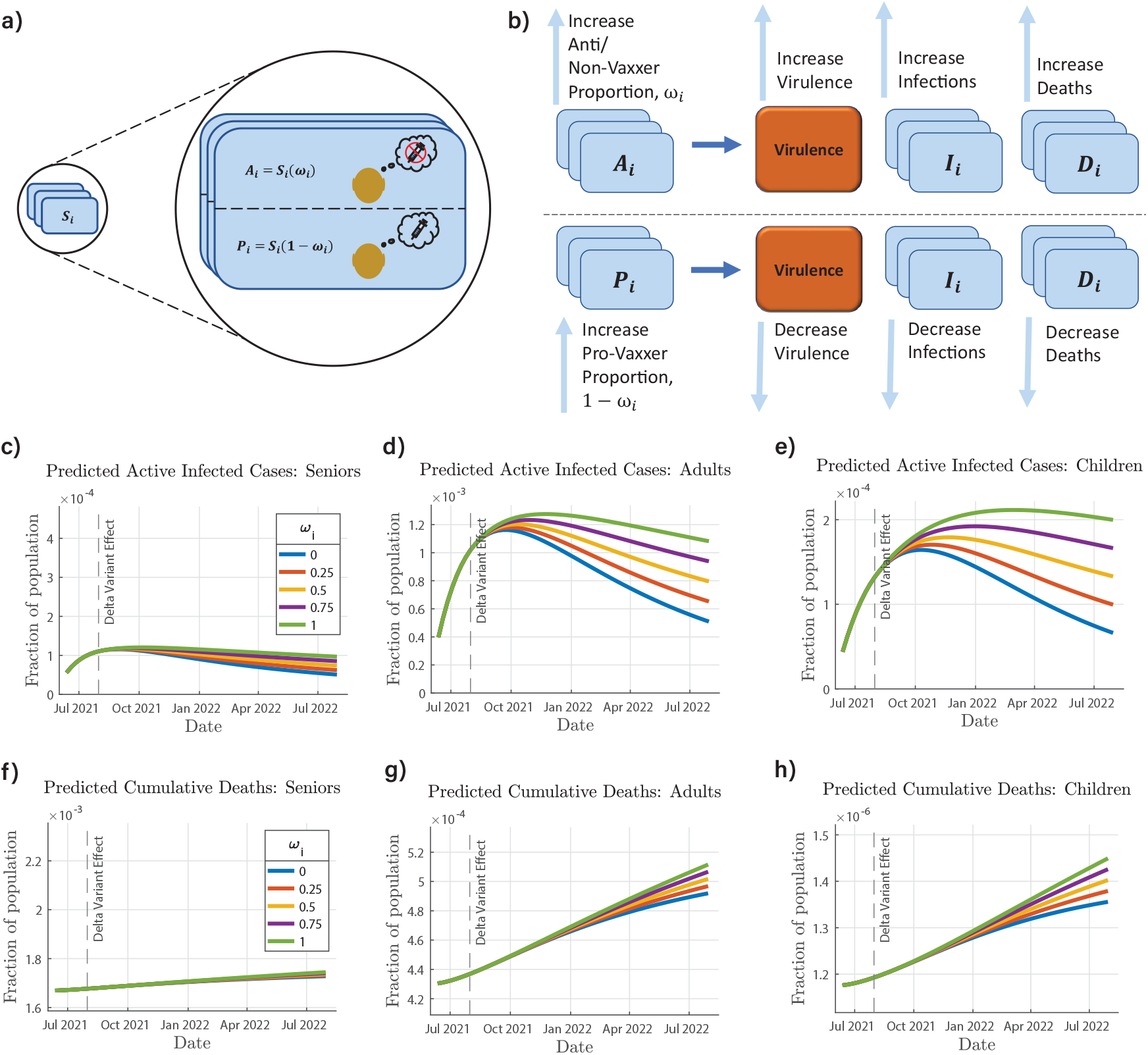
COVID-19 Anti/Non-Vaxxer Effect and the PAIRDV-Virulence Model. (a) The PAIRDV-Virulence model, a variation of the SIRDV-Virulence model, is introduced, in which the susceptible population, *S*_*i*_, is divided into a sub-population that will not be vaccinated, *A*_*i*_, and a sub-population that will be vaccinated, *P*_*i*_. (b) When the proportion of the susceptible population that will not be vaccinated, *ω*_*i*_, increases, the virulence, infections, and deaths increase(c,d,e,f,g,h). The predicted cases and deaths are shown for (c,f) seniors, (d,g) adults, and (e,h) children as a result of simultaneously changing *ω*_*i*_ for each age group.

Dimensionless equations were developed for the PAIRDV-Virulence model in order to conduct the future simulations. Similar to the dimensional model for the PAIRDV-virulence model (see supplementary materials), the dimensionless PAIRDV-virulence model contains one additional differential equation and an additional term in the Infected compartment dynamics compared to the dimensionless SIRDV-virulence model, but contains all the same dimensionless parameters.

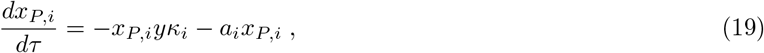

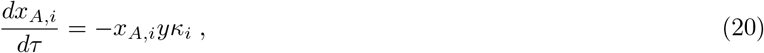

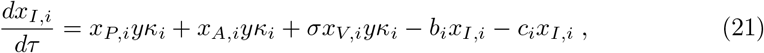

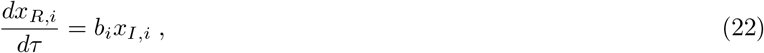

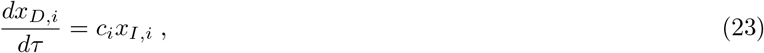

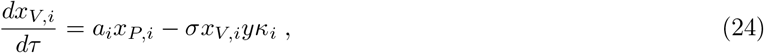

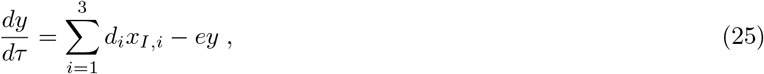

The above differential equations describe the dimensionless behavior of each compartment in the PAIRDV-virulence model, with all the same parameters as the SIRDV-virulence but one additional compartment. For COVID-19 ‘Anti/Non-Vaxxers’, the only way to exit the *x*_*A,i*_ compartment is by COVID-19 infection whereas the COVID-19 ‘Pro Vaxxers’ can exit the *x*_*P,i*_ compartment by either completing their vaccinations or by becoming infected with COVID-19. It is important to note that when *ω*_*i*_ is 0 for all age groups, i, the PAIRDV-Virulence model has the same behavior as the SIRDV-Virulence model, where the COVID-19 ‘Pro Vaxxer’ compartment of the PAIRDV-virulence model acts as the Susceptible compartment of the SIRDV-Virulence model.

As shown in figure 5(b), an increase in the proportion of Anti/Non-Vaxxers will lead to higher virulence and in turn a higher number of infected cases and deaths. The higher the fraction of vaccinated people, the lesser will be the number of deaths and infected cases due to a lower virulence. The future simulations were run in sets, first varying *ω*_*i*_ for each age group while keeping that of the other age groups constant. For these simulations, when *ω*_*i*_ was varied for a single age group, *i*, the dynamics of age group *i* had significant changes, but negligible changes in the dynamics of other age groups were observed (figs. S18-25 in supplementary material). For the second set of simulations, *ω*_*i*_ was varied for all three age groups simultaneously. These results are shown in figures 5c-h. Based on these simulations, an increase in *ω*_*i*_ will result in an increase in cases (figures 5(c-e)) and deaths (figures 5(f-h)) for all three age groups. The proportion of anti-vaxxers affects the children and adult category more than the senior, the reason being a large fraction of seniors has already been vaccinated.

## Discussion

The “future” simulations gave a clear indication that in the case of increased transmission of the mutant variant and possible reduction in the efficacy of the presently administered vaccines, there will be a resurgence in active infected cases and the total number of deaths are also expected to go higher. Among the three age groups considered, the adult and children seem to be affected more than the senior age group. This is justified by the fact a large fraction of senior population has already either been infected or vaccinated, relative to the other two age groups.

In case of unchanged transmission rate of the virus and vaccine efficacy, the peak in active infection is expected to occur sometime in October 2021, whereas if the transmission rate *K*_*i*_ doubles and the vaccine efficacy 1 *− σ* falls to 0.8, the number of active infections at the peak is expected to go higher by about two times and is expected to occur sometime in December 2021. Although there will be a slight increase in total deaths in this worst case scenario, it will not be a significant one, supporting the point that vaccines indeed bring down the mortality rate of COVID-19.

Another interesting observation was that both the active infection and the total deaths seemed to be more strongly dependent on transmission rate of the virus as compared to the vaccine efficacy. This is justified by the structure of our model. In the case of a reduction in vaccine efficacy, the transition from the vaccinated compartment to the infected compartment will increase. However, in case of an increase in the transmission rate of the virus, transition from both the susceptible and vaccinated compartment to the infected compartment will increase.

Thus keeping in mind the predicted resurgence in number of active infected cases in the future for increased transmission rate of the delta variant and reduced efficacy of the vaccine, it was important to develop an optimum vaccine distribution strategy which reduces the peak of active infections and total deaths to the minimum extent possible. The three heat maps (figure 4) were for three different scenarios: priority to seniors, priority to adults, and priority to children. It was seen that the total vaccination and total infected cases seemed to be heavily dependent on both relative dimensionless *a*_1_ (children) and *a*_2_ (adults) as compared to *a*_3_ (seniors). A higher value of relative dimensionless parameter *a*_*i*_ suggests a higher proportion of vaccines being administered to that group. The deaths however were not affected much by *a*_3_ and predominantly affected by *a*_1_ and *a*_2_. These conclusions seem logical as a large fraction of adults and children are still left to be vaccinated and any changes in vaccination rate for these two compartments will affect the overall vaccination and infection. Also the children do not contribute much to the death compartment due to very low mortality rate and hence changes in *a*_1_ won’t affect the total deaths which are heavily dominated by adults and seniors. Since the adult age group consists of the greatest proportion of the US population, any change in vaccination rate for adults will have a drastic effect on the overall dynamics. Thus it can be concluded from the heat maps that prioritizing the adults and children over seniors for vaccination will be a more effective approach for minimizing the future infected cases and deaths and maximizing the fraction of vaccinated population.

The fraction of Anti/Non-Vaxxers had a considerable effect on the active infected cases and total deaths, especially in the children and adult category. For the seniors the effect was not that pronounced, primarily due to the fact that a large fraction of the senior population has either already been infected or vaccinated. For the adult population, a 0.5 fraction of Anti/Non-Vaxxers increases the total death by approximately 6% and causes a delay in reduction rate of the active infected cases. A similar effect is visible in the children age group where an increase of around 3.7% is seen in the total deaths for half of the populated being un-vaccinated. The delay in reduction of active cases in the children age group due to high proportion of Anti/Non-Vaxxers is more pronounced as compared to other age groups.

## Conclusion

With the SARS-CoV-2 virus undergoing mutations at a fast rate, it becomes of immense importance to study the effect of these mutants on the transmissibility rate and vaccine dynamics. While different vaccine manufacturers make different claims about the effectiveness against the mutants, the exact effect of these mutants on vaccine efficacy is still not clear [2]. In such a scenario, it becomes important to predict the effect of the variation on the population, considering increased infection transmissibility and decreased vaccine efficacy to predict the worst case scenarios for future in terms of active infections and total deaths. The predictions of our studies not only help to predict the peak time of infection and help the healthcare system to prepare accordingly but also assist in devising the optimum vaccination strategy, prioritizing the children and adult age groups, which seem to be affected the most by the delta variant.

Along with the introduction of the new variant, another aspect of primary concern is that a certain fraction of the population is not willing to get vaccinated. As the simulations results indicate, the larger the proportion of Anti/Non-Vaxxers, the more time it will take for the number of active infections to come down. Even the number of deaths can increase considerably in the adult and children population if a large fraction is not vaccinated. Thus it remains of primary importance to vaccinate a large fraction of the population in a short time to prevent any resurgence in the near future. The novelty of the study lies in two parts: First, it identifies the age groups which are likely to be worst affected by the delta variant and suggests the optimum vaccine distribution strategy. Secondly, it re-emphasizes the fact that a large fraction of unvaccinated people has catastrophic effects and can cause recurrence of the pandemic. For future studies, it will be interesting to include the dynamics involved with the mutation of the SARS-CoV-2 virus and study how the changing mutation rate can affect the present vaccines being administered. Also, it will be beneficial to identify the specific geographical regions which are likely to get affected the most by the delta variant, taking into account the age distribution in that state and the status of vaccination. These studies will help the health care system to prepare well in advance.

## Data Availability

The data and code that support the findings of this study are available from the 469
corresponding authors. For infection data, COVID-19 weekly cases by age group were 470
collected from the CDC Data Tracker's \COVID-19 Weekly Cases per 100,000 471
Population by Age, Race/Ethnicity, and Sex" data visualization at 472
https://covid.cdc.gov/covid-data-tracker/#demographicsovertime. For each week, data 473
were manually collected for each age group and stored in a csv file. COVID-19 weekly 474
deaths by age group were collected from the CDC's dataset, \Provisional COVID-19 475
Deaths by Week, Sex, and Age" at https://data.cdc.gov/NCHS/Provisional-COVID-19- 476
Deaths-by-Week-Sex-and-Age/vsak-wrfu. The daily cumulative completed vaccinations 477
and daily cumulative administered vaccinations were collected from the CDC's dataset, 478
\COVID-19 Vaccinations in the United States, Jurisdiction" at 479
https://data.cdc.gov/Vaccinations/COVID-19-Vaccinations-in-the-United-States- 480
Jurisdi/unsk-b7fc. Population data for the United States, including total population 481
and age distribution, were collected from the United States Census's \U.S. and World 482
Population Clock" at https://www.census.gov/popclock/.

## Data Availability Statement

The data and code that support the findings of this study are available from the corresponding authors. For infection data, COVID-19 weekly cases by age group were collected from the CDC Data Tracker’s “COVID-19 Weekly Cases per 100,000 Population by Age, Race/Ethnicity, and Sex” data visualization at https://covid.cdc.gov/covid-data-tracker/#demographicsovertime. For each week, data were manually collected for each age group and stored in a csv file. COVID-19 weekly deaths by age group were collected from the CDC’s dataset, “Provisional COVID-19 Deaths by Week, Sex, and Age” at https://data.cdc.gov/NCHS/Provisional-COVID-19-Deaths-by-Week-Sex-and-Age/vsak-wrfu. The daily cumulative completed vaccinations and daily cumulative administered vaccinations were collected from the CDC’s dataset, “COVID-19 Vaccinations in the United States, Jurisdiction” at https://data.cdc.gov/Vaccinations/COVID-19-Vaccinations-in-the-United-States-Jurisdi/unsk-b7fc. Population data for the United States, including total population and age distribution, were collected from the United States Census’s “U.S. and World Population Clock” at https://www.census.gov/popclock/.

## Supporting information

**Fig. S1 Processed CDC case data used for data fitting**.

**Fig. S2 Processed CDC death data used for data fitting**.

**Fig. S3 Processed CDC vaccination data used for data fitting**.

**Fig. S4 Fitted model against completed vaccination data**.

**Fig. S5 Vaccination parameter validation**. a) New weekly completed vaccinations for the fitted time period. b) Fitted model vaccinations with parameter constraint.

**Fig. S6 Model Fitting: Cumulative Cases**. Simulations from mean parameters shown for a) time period 1, b) time period 2, c) time period 3, and d) time period 4.

**Fig. S7 Model Fitting: Cumulative Cases**. Simulations from inter-quartile range of parameters shown for a) time period 1, b) time period 2, c) time period 3, and d) time period 4.

**Fig. S8 Model Fitting: Cumulative Deaths**. Simulations from mean parameters shown for a) time period 1, b) time period 2, c) time period 3, and d) time period 4.

**Fig. S9 Model Fitting: Cumulative Deaths**. Simulations from inter-quartile range of parameters shown for a) time period 1, b) time period 2, c) time period 3, and d) time period 4.

**Fig. S10 Model Fitting: Cumulative Completed Vaccinations**. Simulations from mean parameters shown for a) time period 1, b) time period 2, c) time period 3, and d) time period 4.

**Fig. S11 Model Fitting: Cumulative Completed Vaccinations**. Simulations from inter-quartile range of parameters shown for a) time period 1, b) time period 2, c) time period 3, and d) time period 4.

**Fig. S12 Simulated Active Infections for Simulation Case 1: Effect of Mutation on COVID-19 Transmission**. Variation of transmissibility, *K*, with vaccine inefficacy, *σ* = 0.20.

**Fig. S13 Simulated Deaths for Simulation Case 1: Effect of Mutation on COVID-19 Transmission**. Variation of transmissibility, *K*, with vaccine inefficacy, *σ* = 0.20.

**Fig. S14 Simulated Active Infections for Simulation Case 1: Effect of Mutation on COVID-19 Transmission**. Variation of vaccine inefficacy, *σ* = 0.20, with transmissibility, *K*_*new*_*/K* = 1.

**Fig. S15 Simulated Deaths for Simulation Case 1: Effect of Mutation on COVID-19 Transmission**. Variation of vaccine inefficacy, *σ* = 0.20, with transmissibility, *K*_*new*_*/K* = 1.

**Fig. S16 Simulated Active Infections for Simulation Case 1: Effect of Mutation on COVID-19 Transmission**. Variation of vaccine inefficacy, *σ* = 0.20, with transmissibility, *K*_*new*_*/K* = 2.

**Fig. S17 Simulated Deaths for Simulation Case 1: Effect of Mutation on COVID-19 Transmission**. Variation of vaccine inefficacy, *σ* = 0.20, with transmissibility, *K*_*new*_*/K* = 2.

**Fig. S18 Simulated Active Infections for Simulation Case 3: Effect of Anti/Non-Vaxxers on COVID-19 Tranmission**. Variation of adult Anti/Non-Vaxxer proportion, *ω*_2_, with *ω*_1_ = 0 (children) and *ω*_3_ = 0 (seniors).

**Fig. S19 Simulated Deaths for Simulation Case 3: Effect of Anti/Non-Vaxxers on COVID-19 Tranmission**. Variation of adult Anti/Non-Vaxxer proportion, *ω*_2_, with *ω*_1_ = 0 (children) and *ω*_3_ = 0 (seniors).

**Fig. S20 Simulated Active Infections for Simulation Case 3: Effect of Anti/Non-Vaxxers on COVID-19 Tranmission**. Variation of adult Anti/Non-Vaxxer proportion, *ω*_2_, with *ω*_1_ = 1 (children) and *ω*_3_ = 1 (seniors).

**Fig. S21 Simulated Deaths for Simulation Case 3: Effect of Anti/Non-Vaxxers on COVID-19 Tranmission**. Variation of adult Anti/Non-Vaxxer proportion, *ω*_2_, with *ω*_1_ = 1 (children) and *ω*_3_ = 1 (seniors).

**Fig. S22 Simulated Active Infections for Simulation Case 3: Effect of Anti/Non-Vaxxers on COVID-19 Tranmission**. Variation of children Anti/Non-Vaxxer proportion, *ω*_1_, with *ω*_2_ = 1 (adults) and *ω*_3_ = 1 (seniors).

**Fig. S23 Simulated Deaths for Simulation Case 3: Effect of Anti/Non-Vaxxers on COVID-19 Tranmission**. Variation of children Anti/Non-Vaxxer proportion, *ω*_1_, with *ω*_2_ = 1 (adults) and *ω*_3_ = 1 (seniors).

**Fig. S24 Simulated Active Infections for Simulation Case 3: Effect of Anti/Non-Vaxxers on COVID-19 Tranmission**. Variation of senior Anti/Non-Vaxxer proportion, *ω*_3_, with *ω*_1_ = 1 (children) and *ω*_2_ = 1 (adults).

**Fig. S25 Simulated Deaths for Simulation Case 3: Effect of Anti/Non-Vaxxers on COVID-19 Tranmission**. Variation of senior Anti/Non-Vaxxer proportion, *ω*_3_, with *ω*_1_ = 1 (children) and *ω*_2_ = 1 (adults).

**Table S1. Definition of dimensional variables for SIRDV-Virulence Model**.

## Acknowledgments

The authors gratefully acknowledge the Rising Senior program funded by Professor Sangtae Kim, Jay and Cynthia Ihlenfeld Head of Chemical Engineering and Distinguished Professor at Purdue University.

## Author Contributions

Conceptualization: Doraiswami Ramkrishna, Shiyan Wang.

Data curation: Samuel Heath, Jyotirmoy Roy.

Formal analysis: Jyotirmoy Roy, Samuel Heath.

Funding acquisition: Doraiswami Ramkrishna, Shiyan Wang.

Investigation: Jyotirmoy Roy, Samuel Heath, Doraiswami Ramkrishna, Shiyan Wang.

Methodology: Samuel Heath, Jyotirmoy Roy, Shiyan Wang, Doraiswami Ramkrishna.

Project administration: Doraiswami Ramkrishna, Shiyan Wang.

Resources: Jyotirmoy Roy, Samuel Heath. Software: Samuel Heath, Jyotirmoy Roy.

Supervision: Doraiswami Ramkrishna, Shiyan Wang.

Validation: Jyotirmoy Roy, Samuel Heath, Shiyan Wang.

Visualization: Samuel Heath, Jyotirmoy Roy, Shiyan Wang.

Writing – original draft: Jyotirmoy Roy, Samuel Heath, Shiyan Wang.

Writing – review & editing: Doraiswami Ramkrishna, Shiyan Wang.

## Notes

### Competing Interest Statement

The authors have declared no competing interest.

### Funding Statement

No funding.

