## Supplementary info for "Modeling of COVID-19 Transmission Dynamics on US Population: Inter-transfer Infection in Age Groups, Mutant Variants and Vaccination Strategies"

(Dated: 25 September 2021)

---

[a)] †denotes equal contribution

[b)] Present Address: Department of Mechanical Engineering, Massachusetts Institute of Technology, Cambridge, MA 02139

### I. DYNAMIC MODEL

Our SIRDVV model consisted of 6 compartments: Susceptible, Infected, Recovered, Dead, Vaccinated, and virulence. The relations between them are as follows:

$$\frac{dS_i}{dt} = -\mathscr{V} S_i \beta_i - \alpha_i S_i \,, \tag{1}$$

$$\frac{dI_i}{dt} = \mathscr{V} S_i \beta_i + \mathscr{V} V_i \sigma \beta_i - (1-\mu_i)\gamma_i I_i - \mu_i \rho_i I_i \,, \tag{2}$$

$$\frac{dR_i}{dt} = (1-\mu_i)\gamma_i I_i \,, \tag{3}$$

$$\frac{dD_i}{dt} = \mu_i \rho_i I_i \,, \tag{4}$$

$$\frac{dV_i}{dt} = \alpha_i S_i - \mathscr{V} V_i \sigma \beta_i \,, \tag{5}$$

$$\frac{d\mathscr{V}}{dt} = \sum_{i=1}^{3} (k_{v,i} I_i) - k_v^{'} \mathscr{V} \,, \tag{6}$$

where the variable definitions are defined in Table S1.

### II. DATA PROCESSING PROCEDURE

We consider three different age groups for analysis: children (12-17 years), adults (18-64 years), and seniors (65 years and older).

#### A. COVID-19 Infection Data

The COVID-19 weekly cases data from a CDC visualization of weekly cases by age group were provided with the following age group divisions: 0-4, 5-11, 12-15, 16-17, 18-29, 30-39, 40-49, 50-64, 65-74, and 75 years and older. Only ages 12 and older were considered in this analysis to maintain uniform age groups as those given in the vaccination data set. Since the weekly cases provided by the CDC were given in terms of "per 100,000 Population by Age," the imported data were first adjusted to reflect the total new weekly cases among each age group. Once the data for each age group were adjusted to a weekly new cases basis, the weekly new cases from smaller age groups were summed to create data sets showing the new weekly cases for the three age groups considered in this study: 12-17 years old, 18-64 years old, and 65 years and older. These adjusted data sets were then used to fit the derived model.

| Dimensional Variables | | |
|---|---|---|
| Variable | Significance | Units |
| $S_i$ | *Susceptible* | *Number of people* |
| $I_i$ | *Infected* | *Number of people* |
| $R_i$ | *Recovered* | *Number of people* |
| $D_i$ | *Dead* | *Number of people* |
| $V_i$ | *Vaccinated* | *Number of people* |
| $\mathscr{V}$ | *Virulence* | *?* |
| N | *Total Population* | *Number of people* |
| $v$ | *Viral Population* | *Viral load* |
| $\alpha_i$ | *Vaccination Rate* | *1/time* |
| $\beta_i$ | *Normalised Transmission rate* | *1/(viral load x time)* |
| $\gamma_i$ | *Reciprocal of average recovery time* | *1/time* |
| $\rho_i$ | *Reciprocal of average mortality time* | *1/time* |
| $\mu_i$ | *Mortality Probability* | *Dimensionless* |
| $\sigma_i$ | *Vaccine inefficacy* | *Dimensionless* |
| $k_{v,i}$ | *Viral growth in age group i* | *Viral load/(number of people x time)* |
| $k_v^{'}$ | *Viral death constant* | *1/time* |

TABLE S1. Definition of dimensional variable for SIRDVV model

### B. COVID-19 Mortality Data

COVID-19 death data were imported directly from a CSV file provided by the CDC. These data were given in weekly total new COVID-19 deaths by sex and by age group. The death data were provided with the following age group divisions: 0-1, 1-4, 5-14, 15-24, 25-24, 35-44, 45-54, 55-64, 65-74, 75-84, and 85 years and older. First, the data were filtered to only use data for all sexes. Since age group divisions in this data set were not consistent with those of the three age groups considered in this study (12-17, 18-64, and 65+), the data were processed to approximate the deaths among new age group divisions. For example, in the 15-24 age group, it was estimated that each age in the age group range experienced the same number of deaths. This assumption was validated by a relatively flat age group distribution in this age range. Specifically, it was assumed that 30 percent of the deaths in the 15-24 age group range would happen in the 15-17 age group range and 70 percent of the deaths in the 15-24 age group range would be experienced in the 18-24 age group range. Thus, the 15-24 age group was split into two smaller age groups (15-17 and 18-24) to reflect the age groups chosen for the study. In the same manner, the 5-14 age group range was split into two age groups: 5-11 and 12-14. Since a small number of deaths have been experienced in the younger, modified age groups (relative to the adult and senior age groups), any error in the assumption of an even death distribution will not have a significant effect on the model results. Once the appropriate age groups were created, the weekly new deaths from smaller age groups were summed to create data sets containing the new weekly deaths for the three age groups considered in this study: 12-17 years old, 18-64 years old, and 65 years and older. These adjusted, processed data sets were then used to fit the derived model.

### C. COVID-19 Vaccination Data

COVID-19 vaccination data were imported directly from a CSV file provided by the CDC. Because data were provided for each state in the United States and for the United States as a whole, the data were first filtered to only import vaccination data for the United States as a whole. For simplification of the model analysis in this study, it was assumed that once an individual had received their final vaccination (the first of a one-dose vaccination or the second of a two-dose vaccination), they would move into the Vaccinated compartment. Since the data from the CDC only had completed vaccinations available starting after March 6, 2021, different strategies were used for sorting and processing the vaccination data before and after March 6, 2021, as described below.

Following March 6, 2021, the CDC provided data for total cumulative completed vaccinations on a daily basis for the following age groups: 12 years and older, 18 years and older, and 65 years and older. After importing these data sets, they were adjusted to reflect the following age groups: 12-17, 18-64, and 65 years and older. Next, these data sets were adjusted to change the total daily cumulative completed vaccinations to daily new completed vaccinations. Following this step, for each age group, daily new completed vaccinations were summed up over each week to express the data as weekly new completed vaccinations. These adjusted, processed data sets were then used to fit the derived model.

### D. Vaccination Timeline & Vaccine Types

For the time period up to and including March 6, 2021, the CDC did not provide data on completed vaccinations. Thus, these data were estimated using other available data in the CSV file. Specifically, for the dates from December 13, 2020 to March 6, 2021, the number of total cumulative daily administered vaccines (for all age groups and for the United States) were imported, sorted, and processed to estimate the weekly new completed vaccinations. Because there are no reported vaccinations administered for the children age group (12-17 years old) until May 13, 2021, it is assumed that there are no vaccinations administered to children during the period prior to March 6, 2021. Therefore, completed vaccinations are only estimated for the adult and senior age group. Based on the data from the CDC, there were a negligible amount of Janssen vaccines administered prior to March 6, 2021, with almost all identified vaccines administered during this time period being Moderna and Pfizer. Since the Moderna and Pfizer COVID-19 vaccinations are both 2-dose vaccines[1,2], two major assumptions were made for the data processing: 1) All vaccinations administered during the pre-March 6, 2021 time period were 2 dose vaccinations. 2) The vaccination is considered completed 4 weeks after the first dose is administered (based on the time period between Moderna vaccination doses (1 month)[1], which give a more conservative estimate than the 3-week time period between Pfizer doses[2]). The total cumulative daily administered vaccination vaccines were processed in the following manner. First, the data set was adjusted from cumulative daily administered vaccinations to new daily administered vaccinations. Next, it was adjusted to reflect new weekly administered vaccinations. Following this step, the data were adjusted to estimate weekly administered first doses. Next, a four-week time delay was applied to the data set so that all individuals would be considered as completing their vaccine 4 weeks after the administration of their first dose. Lastly, the processed data for the adult age group and senior age group were each multiplied by a calculated factor so that the cumulative estimated weekly completed vaccines for each age group would accurately reflect the CDC reported numbers starting after March 6, 2021. In short, the data for administered vaccinations was utilized to estimate the distribution of the completed vaccinations for the adult and senior age groups prior to March 6, 2021, and the completed vaccination data (starting after March 6, 2021) was utilized to adjust the values of the estimated pre-March 6 data to achieve a continuous distribution for completed vaccinations during the two time periods.

### E. Data Fitting Procedure

As shown in the dynamic SIRDV-virulence model equations, individuals can enter the death compartment but cannot leave it. Thus, the number of individuals in the Death compartment can be considered as the cumulative total COVID-19 deaths from the beginning of the pandemic to the starting week of the time period of analysis. Thus, the initial value for the Death compartment is the sum of weekly new deaths from the beginning of the death data set to the starting week of analysis.

Since the rate of movement into the Vaccination compartment is significantly larger than the rate of movement out of the Vaccination compartment, the initial value of individuals in the Vaccination compartment is estimated by calculating the cumulative total completed vaccinations from the first date of completed vaccinations until the starting week of analysis. Although this assumption leads to a slight overestimation of initial value for individuals in the Vaccinated compartment, it will have a negligible effect on estimating the fitted parameters and predicting the future dynamics of the pandemic.

Because the case data available were for weekly new cases (and not active infected cases), the weekly new cases data was utilized to estimate the active infected cases in the Infected compartment. The Infected compartment includes the number of individuals that have active infections, with comparable rates of movement into and out of this compartment. Because most infected individuals recover from the virus within 2 weeks[3], the initial value for the Infected compartment is estimated by summing the total new cases for the first week of analysis and the preceding week.

Data were not available for the recovered population, so the death and case data were used to estimate the initial value of individuals in the Recovered compartment at the beginning of the time period of analysis. This value was estimated by summing the weekly new cases from the beginning of the pandemic to the week three weeks before the the initial week of analysis (to exclude those in the Infected compartment) and then subtracting the total cumulative deaths up to and including the initial week of analysis (to exclude those in the Death compartment).

### F. Data Fitting

The raw, normalized data for vaccinations, infected cases and deaths were fitted with the SIRDV-virulence model to obtain the parameters related to transmissibility, vaccination, recovery, mortality, virus multiplication and viral load removal. Rather than fitting weekly cases, weekly deaths, and weekly completed vaccinations, fitting the cumulative cases, deaths, and completed vaccinations resulted in a better fit. The fminsearch function of MATLAB was used for this fitting and the function to be minimized was defined as the summation square of errors for COVID-19 cases, COVID-19 deaths, and completed COVID-19 vaccinations for each age group, with each error term being normalised by the maximum value for the respective set over all time points. For example, when analyzing the vaccination term for adults, the sum of square errors is simply the difference between the actual cumulative vaccinated adults and predicted cumulative vaccinated adults, normalised by the maximum number of actual accumulated vaccinated people, and finally a square of this term. There were 9 such terms for each time point since for each of the three age groups, there are three terms: infected accumulated cases, accumulated deaths, and accumulated vaccinated people.

Before fitting the SIRDV-virulence model, it was necessary to obtain the guess parameters for the fitting. Therefore, a phased approach was used. First, the SIRDV-virulence model was simulated for one age group at a time to obtain the initial guess parameters which produced curves with similar behavior to the actual data. Using these guess values, SIRDV-virulence model was fitted for one age group at a time to obtain parameters for each age group. To do this, the system of ODEs were repeatedly simulated with a new set of guess parameters, obtained from guess parameters multiplied by a random number. The fitted set of parameter was considered to be valid only if all 16 were positive. The first 100 sets of such parameters were considered and the fitted parameters reported were an average over these 100 sets. Once the fitted parameters for each age group were obtained, these were used as guess values for the all age group combined SIRDV-virulence model. Again fitting simulations were run, until the set of first 100 all positive parameters were obtained using the method described for individual age groups. The final reported parameter set is the average of these 100 trials.

### G. PAIRDV-Virulence Model

To simulate ’Anti/Non-Vaxxer’ effect, a modified compartmental model, PAIRDV-virulence, was developed. The equations are shown below

$$\frac{dP_i}{dt} = -\mathcal{V} P_i \beta_i - \alpha_i P_i \,, \tag{7}$$
$$\frac{dA_i}{dt} = -\mathcal{V} A_i \beta_i \,, \tag{8}$$
$$\frac{dI_i}{dt} = \mathcal{V} P_i \beta_i + \mathcal{V} A_i \beta_i + \mathcal{V} V_i \sigma \beta_i - (1-\mu_i)\gamma_i I_i - \mu_i \rho_i I_i \,, \tag{9}$$
$$\frac{dR_i}{dt} = (1-\mu_i)\gamma_i I_i \,, \tag{10}$$
$$\frac{dD_i}{dt} = \mu_i \rho_i I_i \,, \tag{11}$$
$$\frac{dV_i}{dt} = \alpha_i P_i - \mathcal{V} V_i \sigma \beta_i \,, \tag{12}$$
$$\frac{d\mathcal{V}}{dt} = \sum_{i=1}^{3} (k_{v,i} I_i) - k_v^{'} \mathcal{V} \,, \tag{13}$$

There are two major assumptions for the PAIRDV-virulence model: 1) COVID-19 'Pro Vaxxers' and 'Anti/Non-Vaxxers' will have the same likelihood of transmitting the virus (prior to obtaining immunity from a completed vaccination). 2) The only way to move into the Vaccinated compartment is from the COVID-19 'Pro Vaxxer' compartment, $P_i$. In other words, the PARIDV-virulence model considers the vaccination of susceptible COVID-19 'Pro Vaxxers' and not the vaccination of Recovered individuals.

To simulate future predictions with the PAIRDV-virulence compartmental model, parameters from the fourth fitted time period were used, and the $\omega$ value was varied for each age group to see the effect on the future dynamics, mainly how the change in $\omega$ changed the number of cases and deaths in each age group over time.

### H. Supporting figures

### REFERENCES

[1]Moderna, "Moderna covid-19 vaccine fact sheet for recipients and caregivers," https://www.fda.gov/media/144638/download.

[2]Pfizer and BioNTech, "Final$_{e}ua_{f}ullpi_{h}cpfactsheetpfizer-biontechcovid-19vaccine,''$ .

H. H. Publishing, noop If you've been exposed to the coronavirus, https://www.health.harvard.edu/diseases-and-conditions/if-youve-been-exposed-to-the-coronavirus.

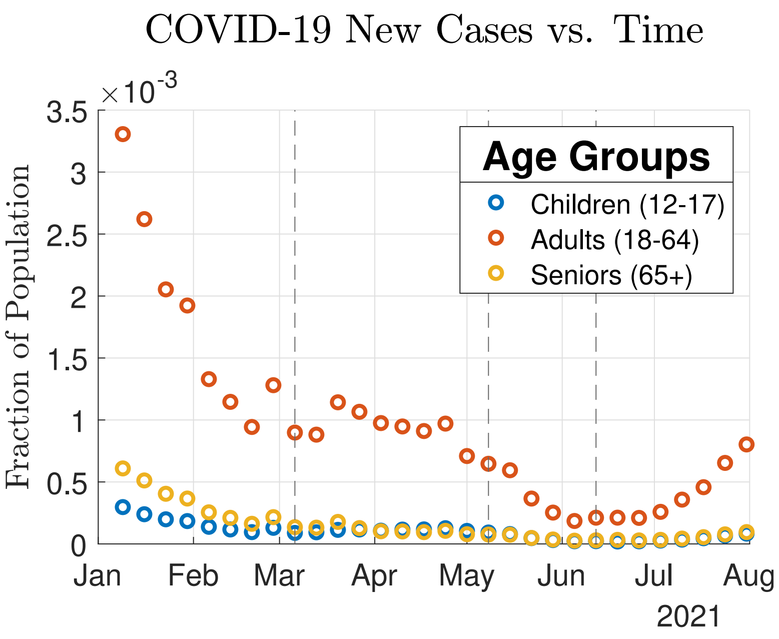


FIG. S1: Processed CDC case data used for data fitting.

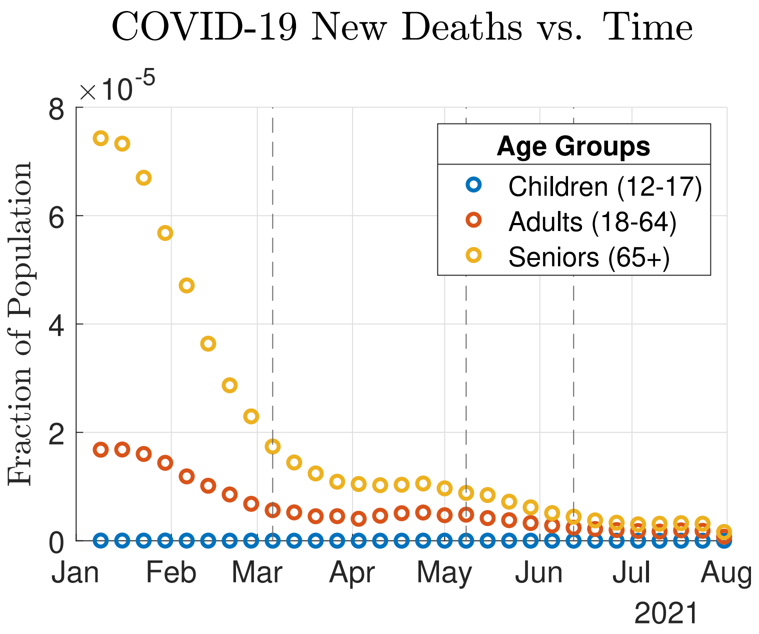


FIG. S2: Processed CDC death data used for data fitting.

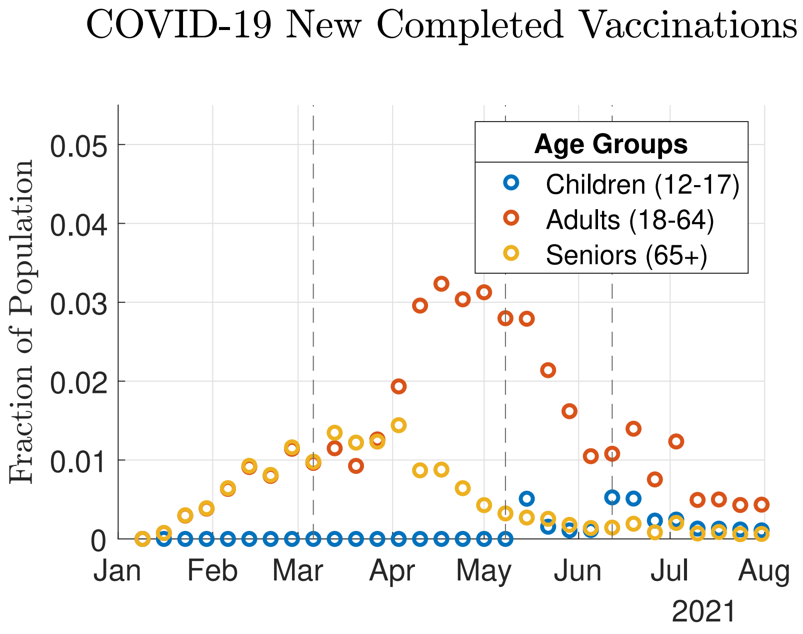


FIG. S3: Processed CDC vaccination data used for data fitting.

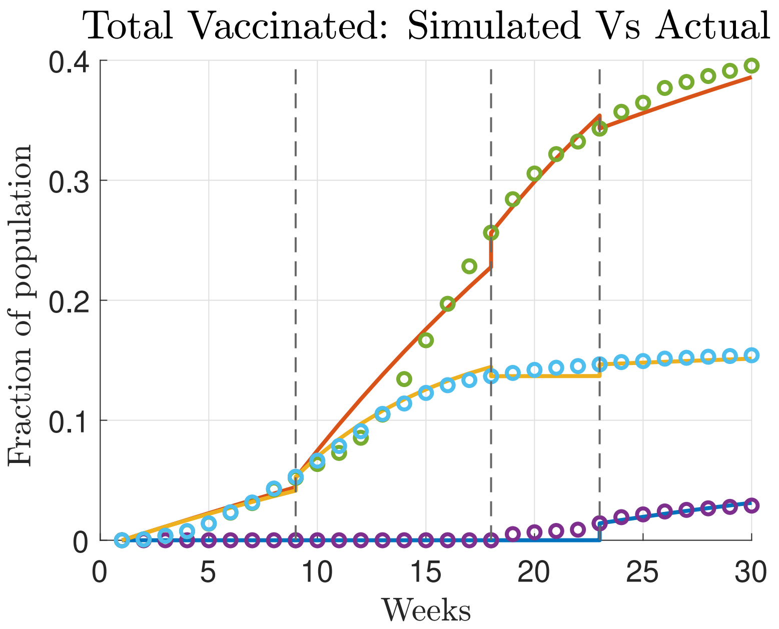


FIG. S4: Fitted model against cumulative completed vaccinations. The adult age group corresponds to the red line and green circles, seniors to yellow line and cyan circles, children to blue line and purple circles.

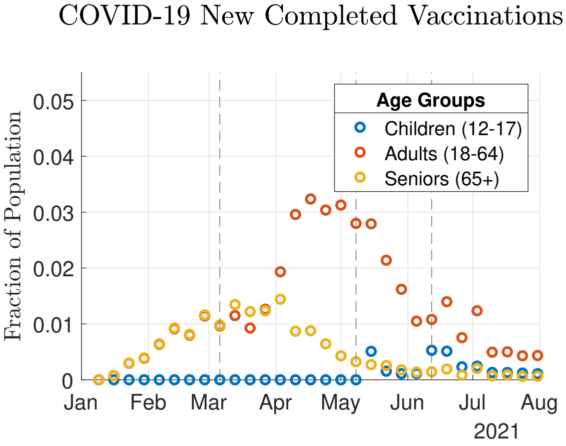


(a) Completed Vaccinations Data

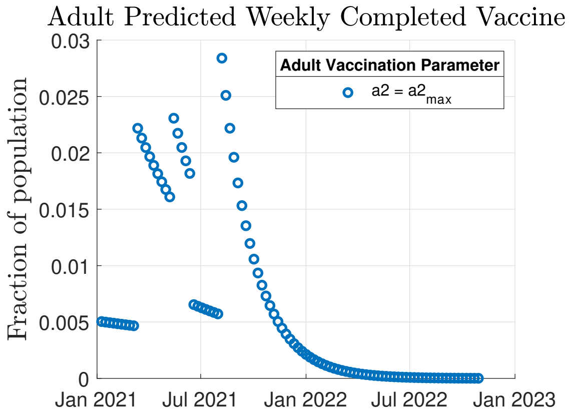


(b) Model of Completed Vaccines

FIG. S5: Vaccination parameter validation. a) New weekly completed vaccinations for the fitted time period. During this time period, 0.032 is the highest adult fraction (relative to the total studied population) of weekly completed vaccinations. b) The fitted model is run to simulate weekly completed vaccinations and calculate the maximum practical vaccination parameter for adults (the highest value that doesn't exceed a weekly completed vaccination rate of 0.032.)

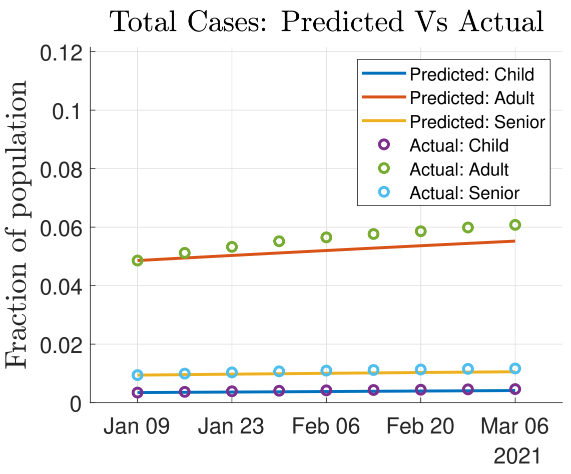


(a) Time Period 1 (1/9/21 - 3/6/21)

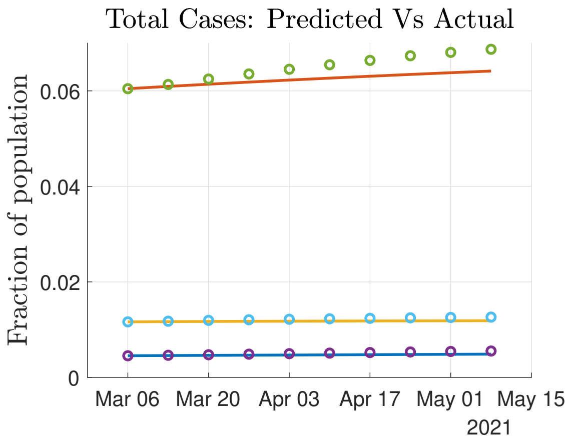


(b) Time Period 2 (3/6/21 - 5/8/21)

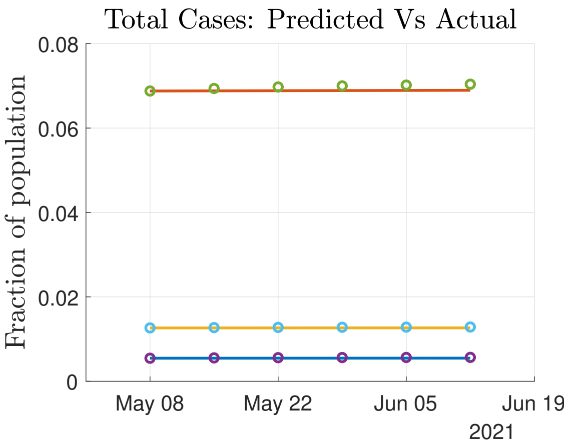


(c) Time Period 3 (5/8/21 - 6/12/21)

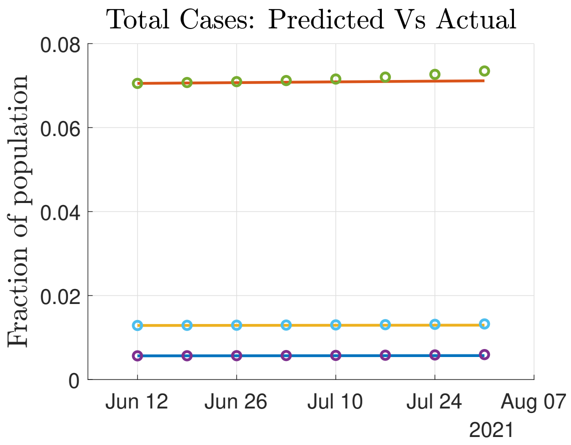


(d) Time Period 4 (6/12/21 - 7/31/21)

FIG. S6: Model Fitting: Cumulative cases fitted to processed data for a) time period 1, b) time period 2, c) time period 3, and d) time period 4. Only mean values shown.

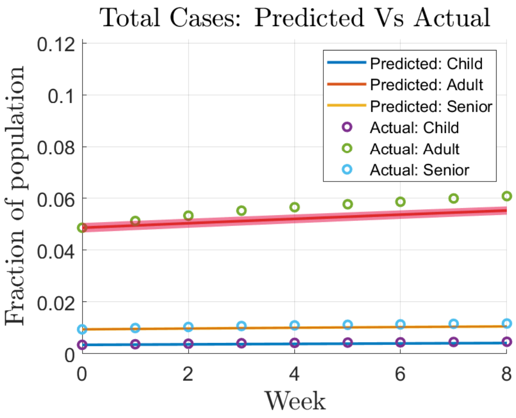


(a) Time Period 1 (1/9/21 - 3/6/21)

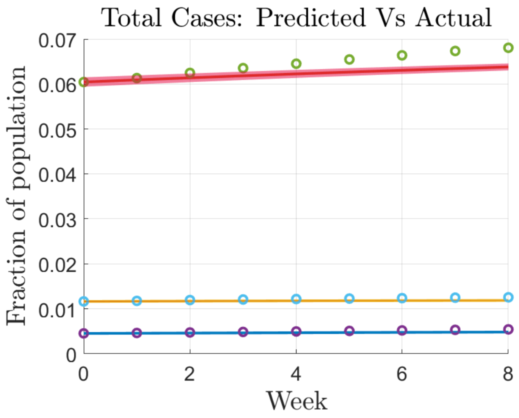


(b) Time Period 2 (3/6/21 - 5/8/21)

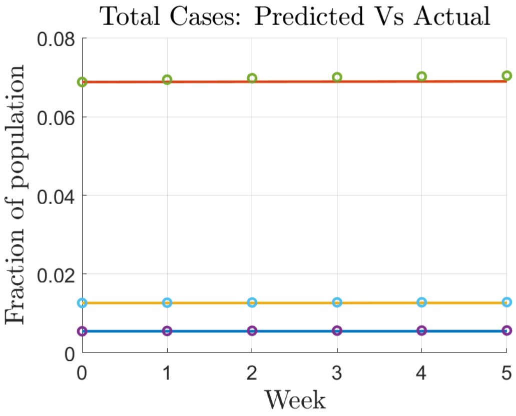


(c) Time Period 3 (5/8/21 - 6/12/21)

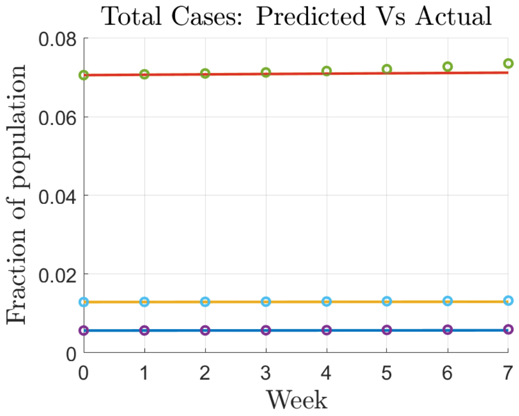


(d) Time Period 4 (6/12/21 - 7/31/21)

FIG. S7: Model Fitting: Cumulative cases fitted to processed data for a) time period 1, b) time period 2, c) time period 3, and d) time period 4. Inter-quartile range of model parameters shown.

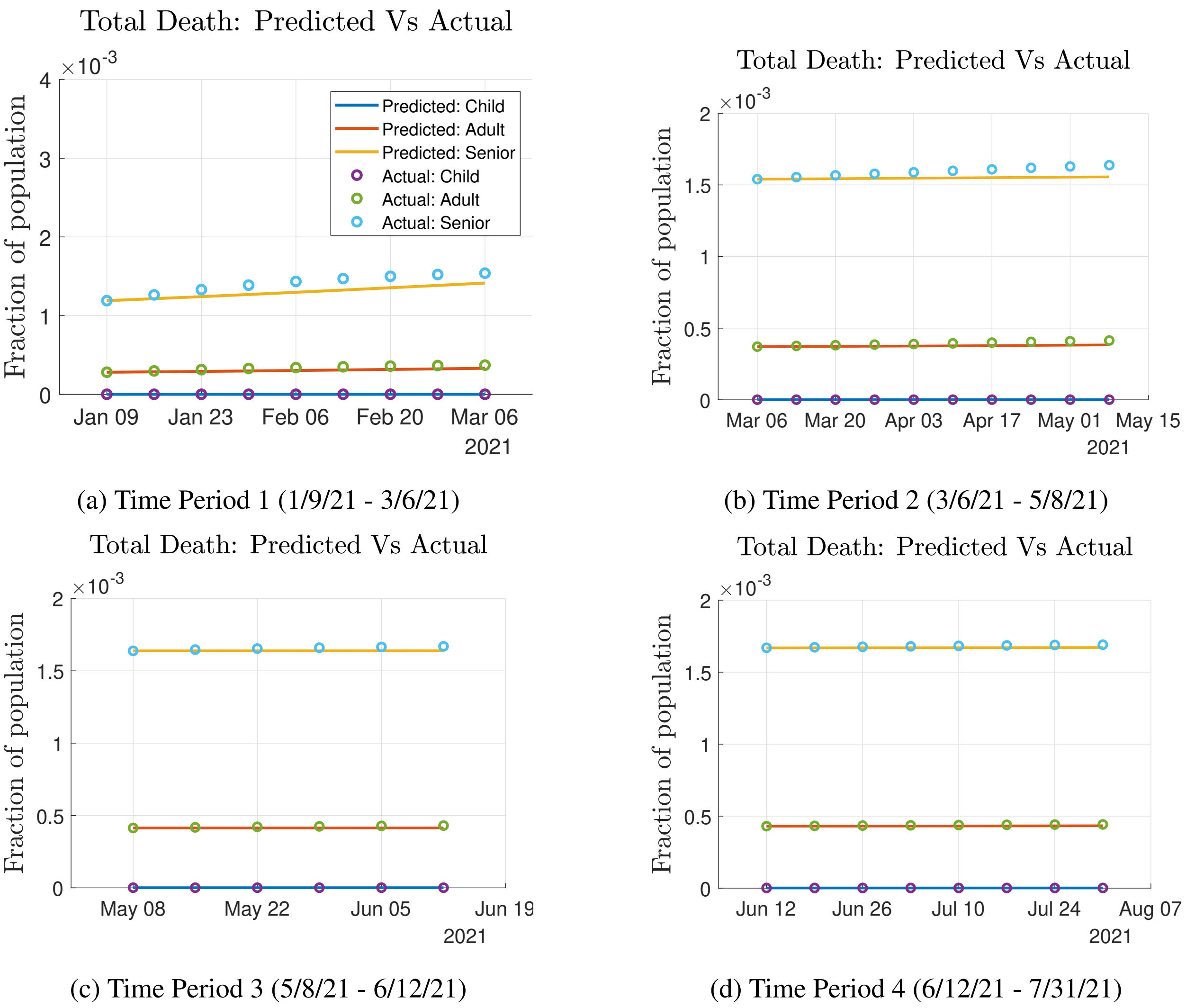


(a) Time Period 1 (1/9/21 - 3/6/21)

(b) Time Period 2 (3/6/21 - 5/8/21)

(c) Time Period 3 (5/8/21 - 6/12/21)

(d) Time Period 4 (6/12/21 - 7/31/21)

FIG. S8: Model Fitting: Cumulative deaths fitted to processed data for a) time period 1, b) time period 2, c) time period 3, and d) time period 4. Only mean values shown.

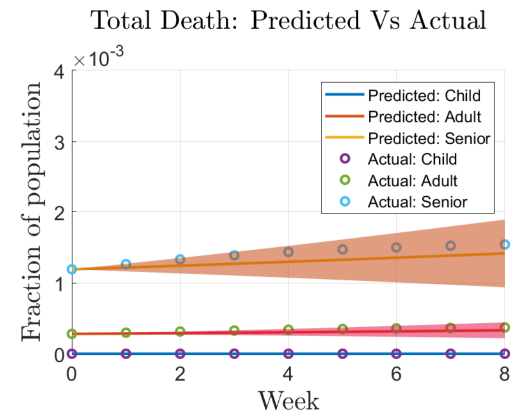


(a) Time Period 1 (1/9/21 - 3/6/21)

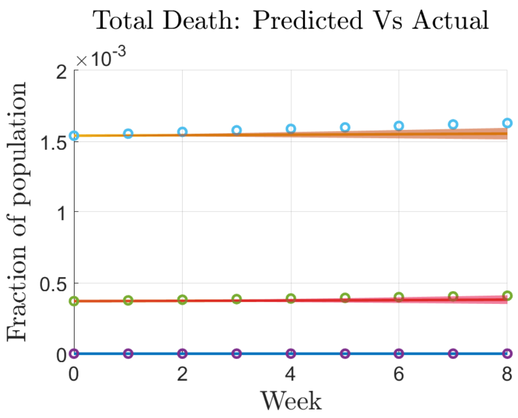


(b) Time Period 2 (3/6/21 - 5/8/21)

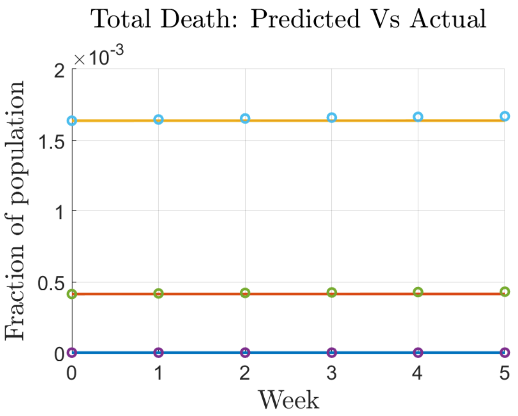


(c) Time Period 3 (5/8/21 - 6/12/21)

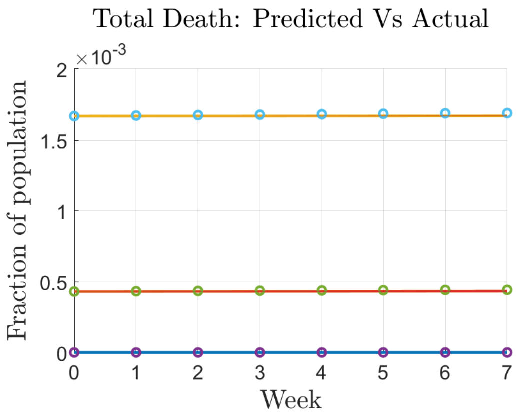


(d) Time Period 4 (6/12/21 - 7/31/21)

FIG. S9: Model Fitting: Cumulative deaths fitted to processed data for a) time period 1, b) time period 2, c) time period 3, and d) time period 4. Inter-quartile range of model parameters shown.

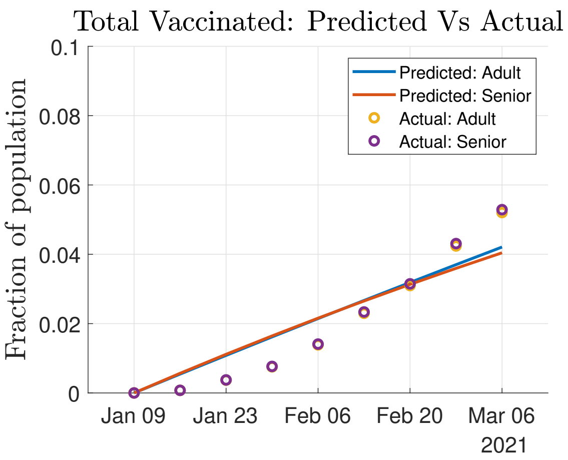


(a) Time Period 1 (1/9/21 - 3/6/21)

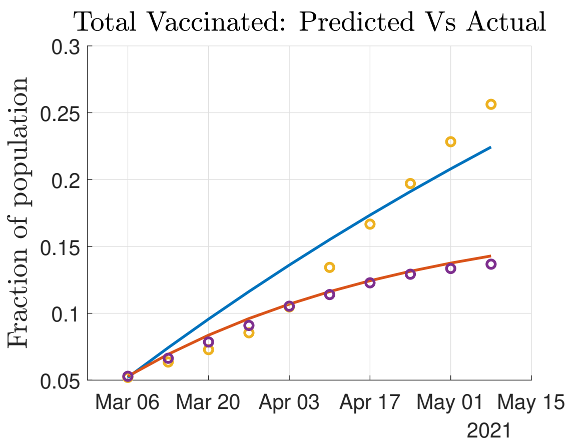


(b) Time Period 2 (3/6/21 - 5/8/21)

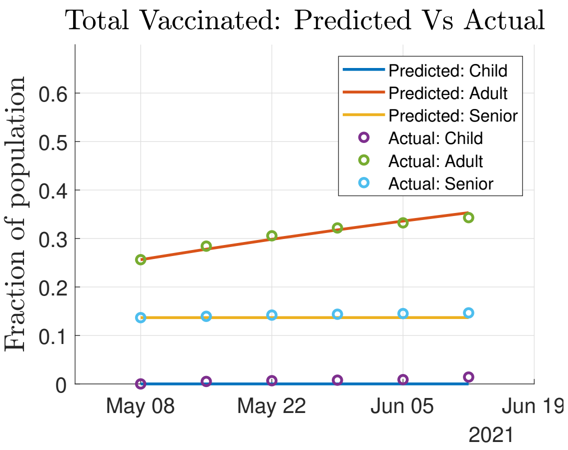


(c) Time Period 3 (5/8/21 - 6/12/21)

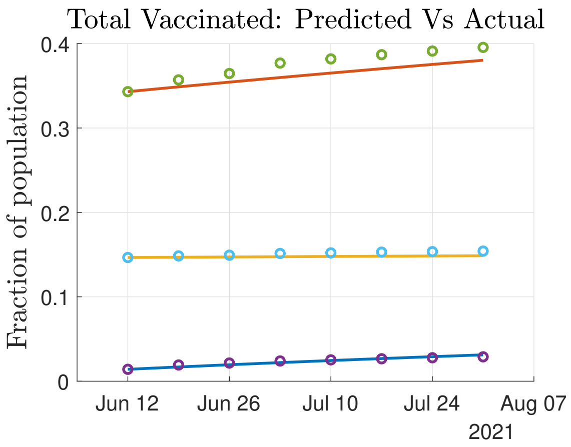


(d) Time Period 4 (6/12/21 - 7/31/21)

FIG. S10: Model Fitting: Cumulative completed vaccinations fitted to processed data for a) time period 1, b) time period 2, c) time period 3, and d) time period 4. Only mean values shown.

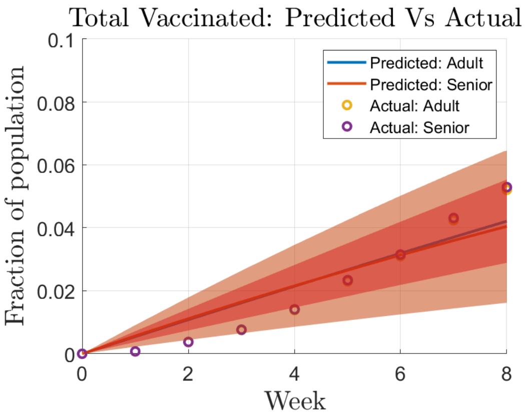


(a) Time Period 1 (1/9/21 - 3/6/21)

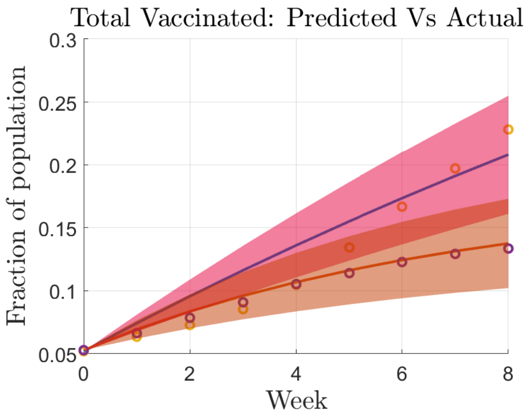


(b) Time Period 2 (3/6/21 - 5/8/21)

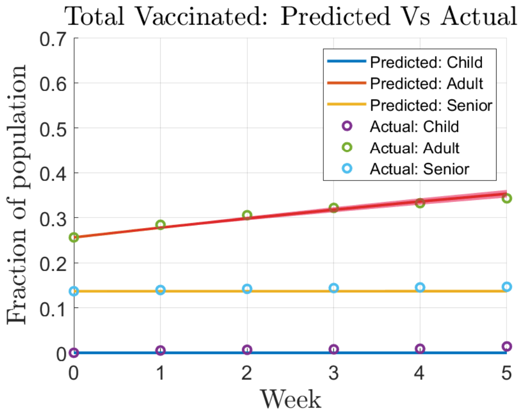


(c) Time Period 3 (5/8/21 - 6/12/21)

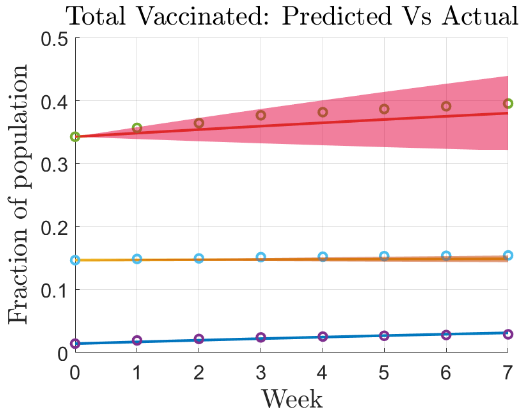


(d) Time Period 4 (6/12/21 - 7/31/21)

FIG. S11: Model Fitting: Cumulative completed vaccinations fitted to processed data for a) time period 1, b) time period 2, c) time period 3, and d) time period 4. Inter-quartile range of model parameters shown.

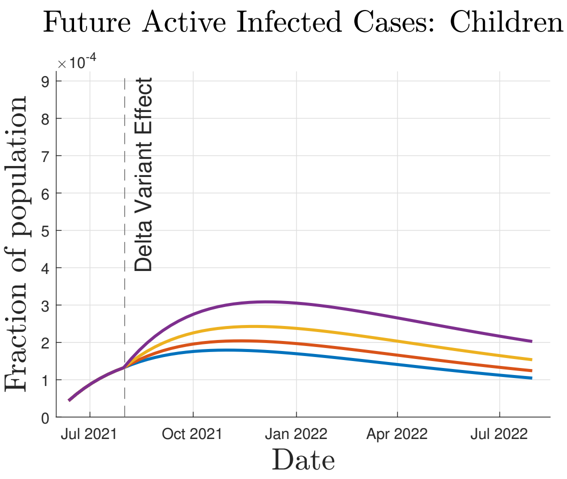


(a) Children

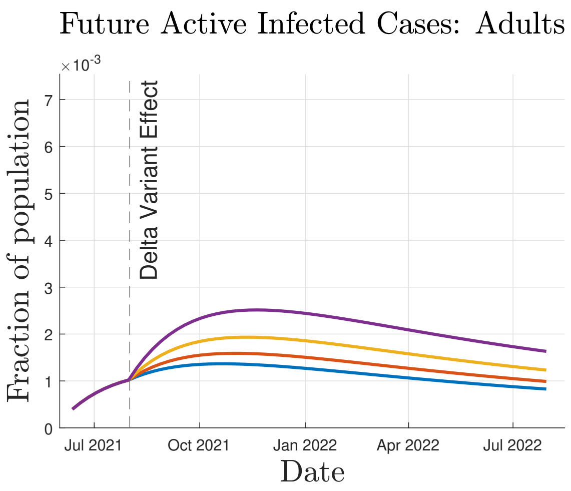


(b) Adults

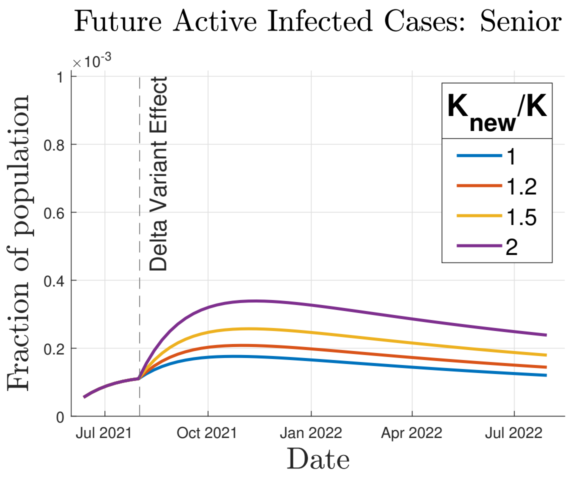


(c) Seniors

FIG. S12: Simulation Case 1: Effect of Mutation on COVID-19 Transmission. Variation of transmissibility parameter, $K$, and vaccine inefficacy, $\sigma = 0.20$. Simulated Active Infections for a) children, b) adults, and c) seniors.

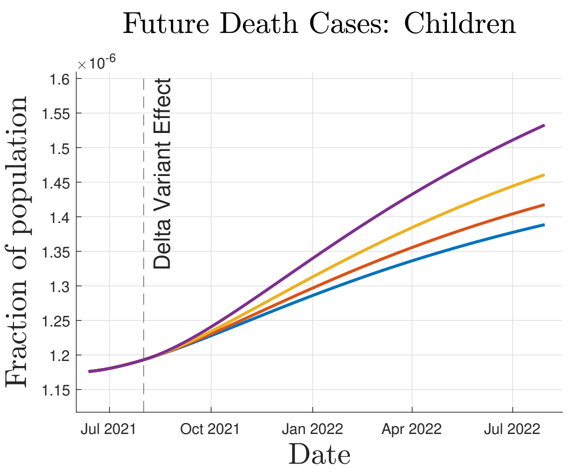


(a) Children

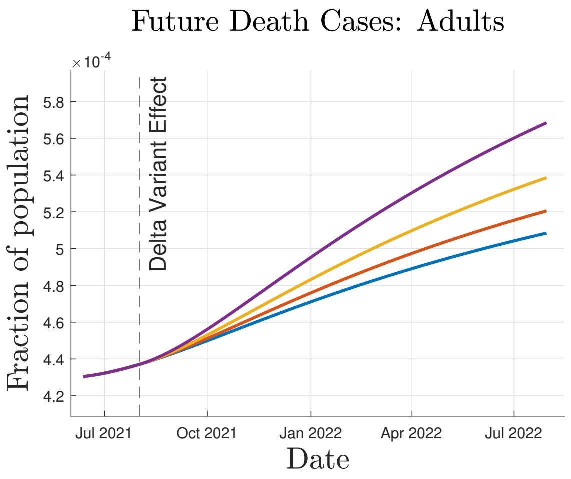


(b) Adults

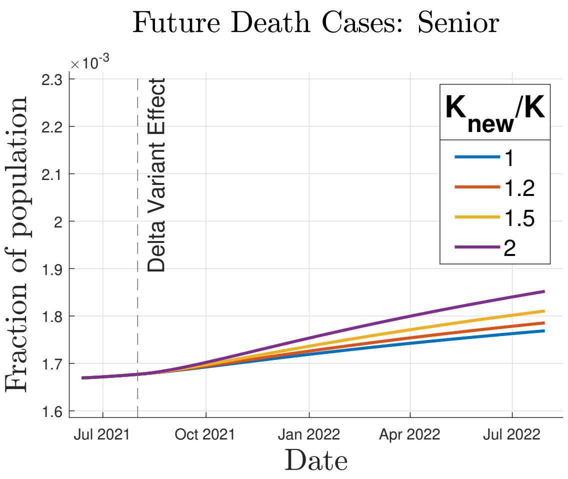


(c) Seniors

FIG. S13: Simulation Case 1: Effect of Mutation on COVID-19 Transmission. Variation of transmissibility parameter, $K$, and vaccine inefficacy, $\sigma = 0.20$. Simulated Deaths for a) children, b) adults, and c) seniors.

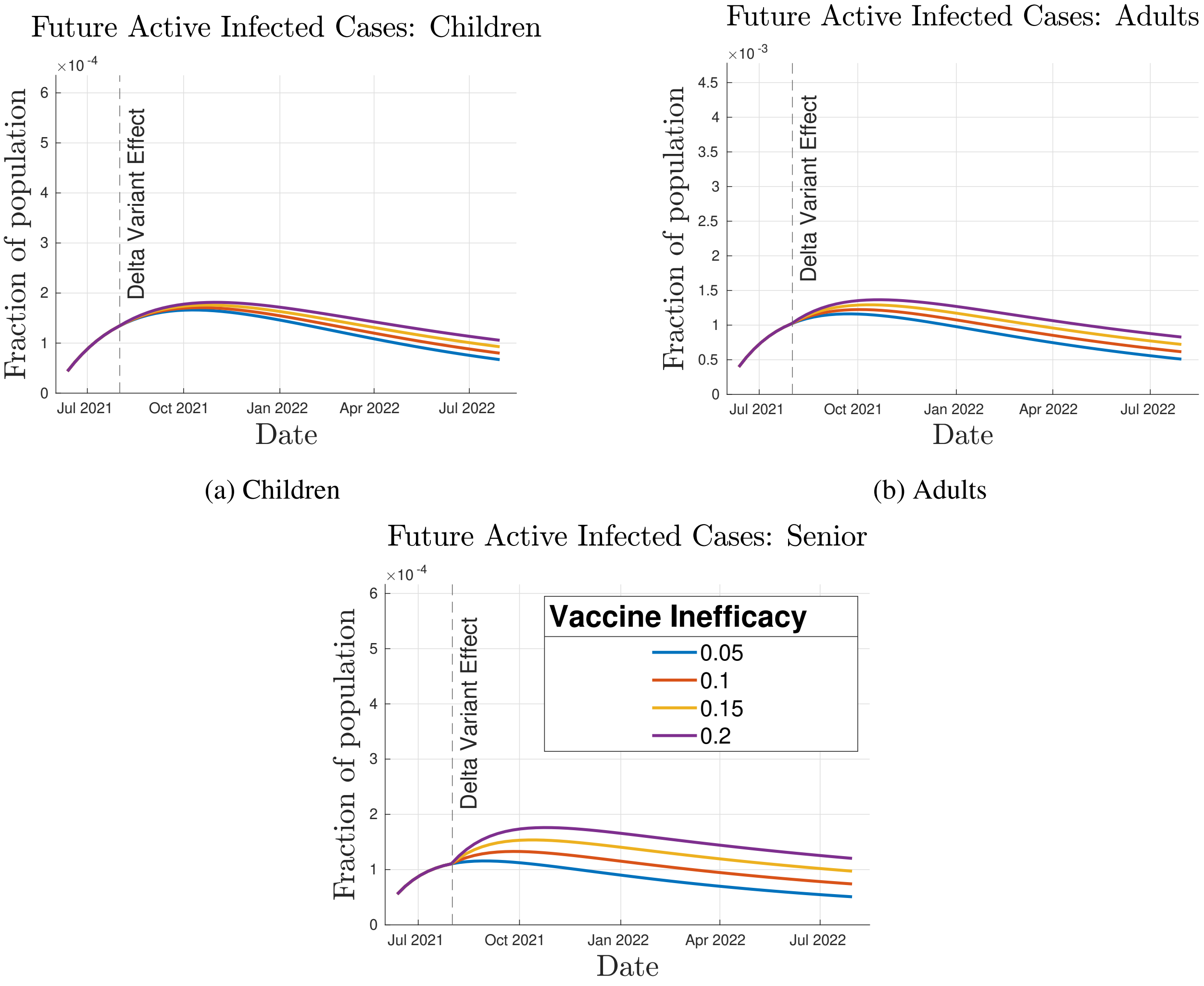


(a) Children

(b) Adults

(c) Seniors

FIG. S14: Simulation Case 1: Effect of Mutation on COVID-19 Transmission. Variation of vaccine inefficacy, $\sigma$, with transmissibility parameter, $K_{new}/K = 1$. Simulated Active Infected Cases for a) children, b) adults, and c) seniors.

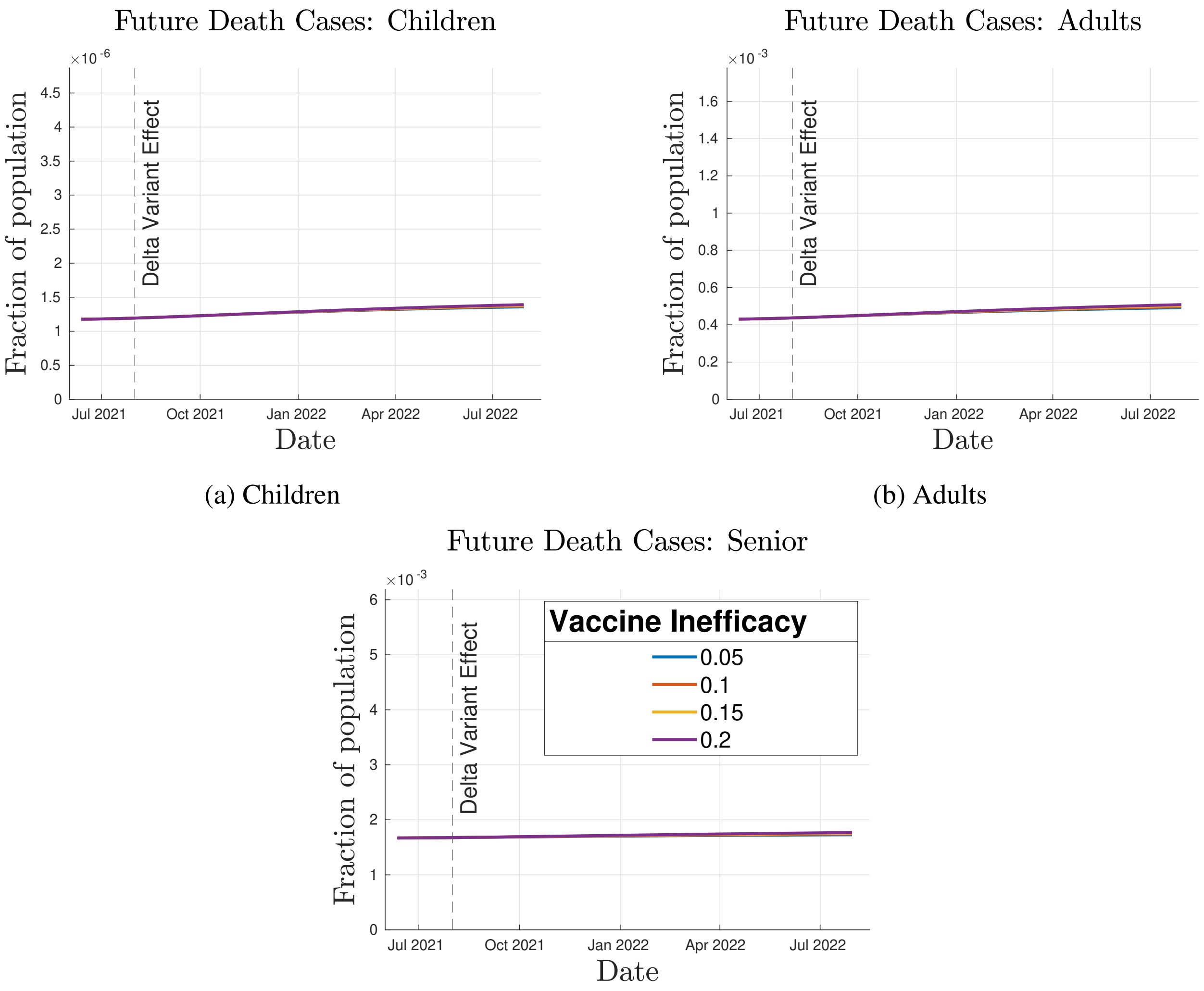


(a) Children

(b) Adults

(c) Seniors

FIG. S15: Simulation Case 1: Effect of Mutation on COVID-19 Transmission. Variation of vaccine inefficacy, $\sigma$, with transmissibility parameter, $K_{new}/K = 1$. Simulated Deaths for a) children, b) adults, and c) seniors.

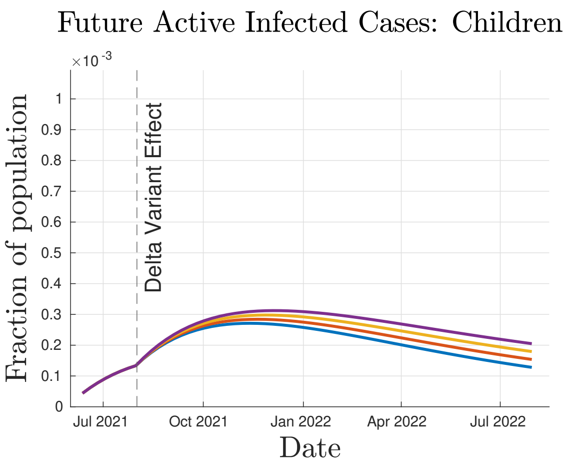


(a) Children

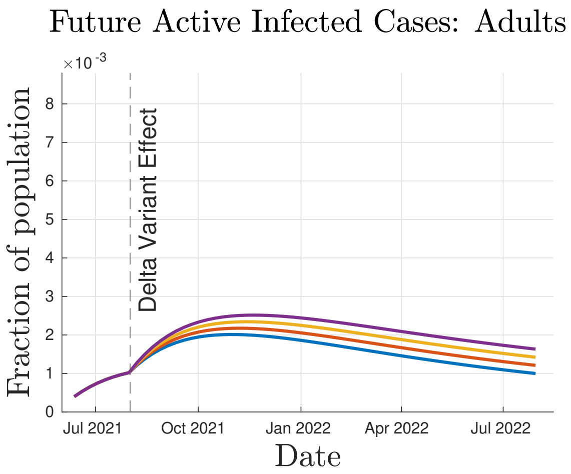


(b) Adults

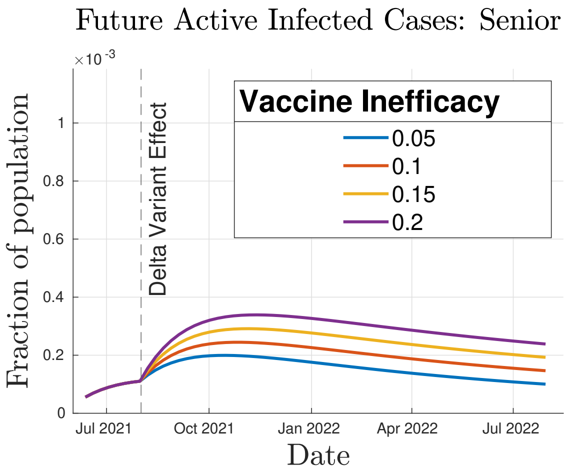


(c) Seniors

FIG. S16: Simulation Case 1: Effect of Mutation on COVID-19 Transmission. Variation of vaccine inefficacy, $\sigma$, with transmissibility parameter, $K_{new}/K = 2$. Simulated Active Infected Cases for a) children, b) adults, and c) seniors.

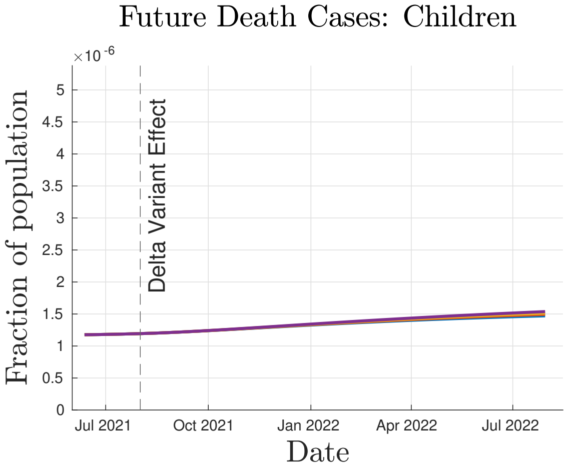


(a) Children

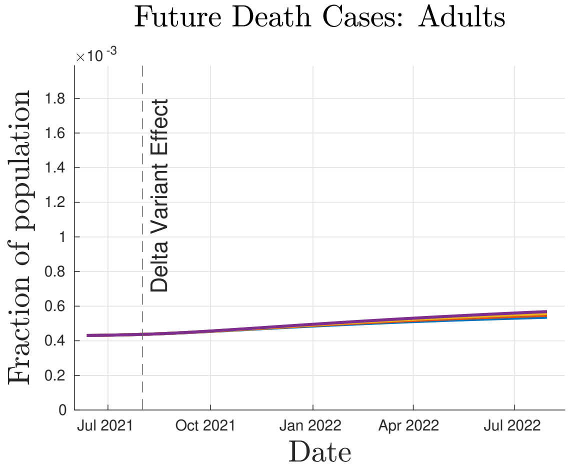


(b) Adults

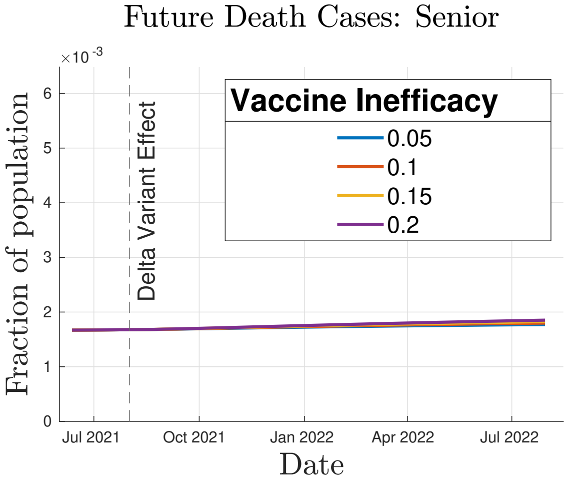


(c) Seniors

FIG. S17: Simulation Case 1: Effect of Mutation on COVID-19 Transmission. Variation of vaccine inefficacy, $\sigma$, with transmissibility parameter, $K_{new}/K = 2$. Simulated Deaths for a) children, b) adults, and c) seniors.

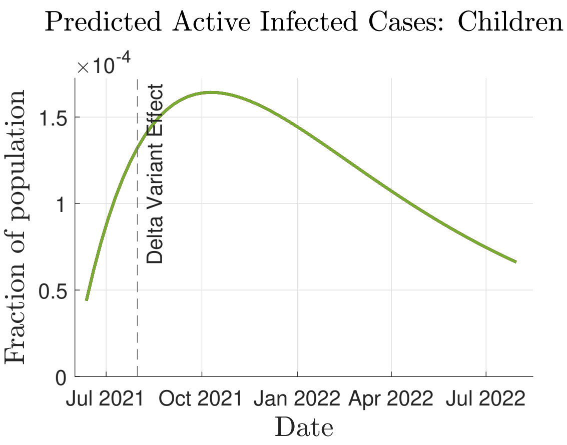


(a) Children

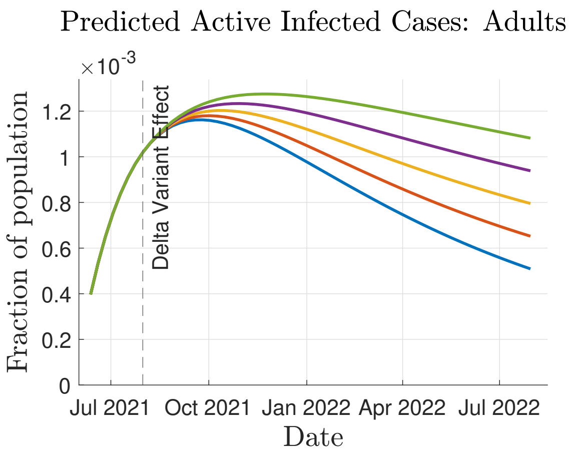


(b) Adults

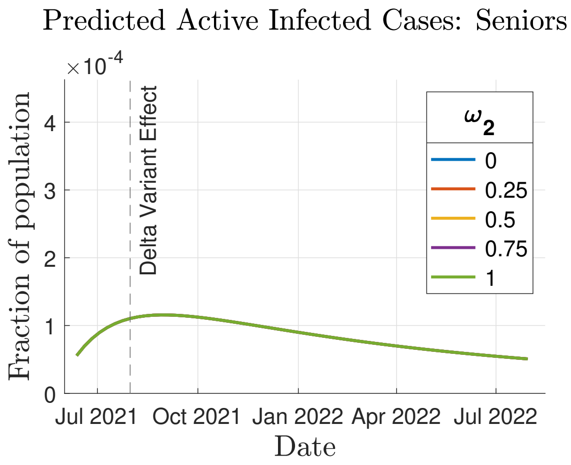


(c) Seniors

FIG. S18: Simulation Case 3: Effect of Anti/Non-Vaxxers on COVID-19 Transmission. Variation of adult Anti/Non-Vaxxer proportion, $\omega_2$, with $\omega_1 = 0$ (children) and $\omega_3 = 0$ (seniors). Vaccine inefficacy, $\sigma = 0.05$, and transmissibility parameter, $K_{new}/K = 1$. Simulated Infections for a) children, b) adults, and c) seniors.

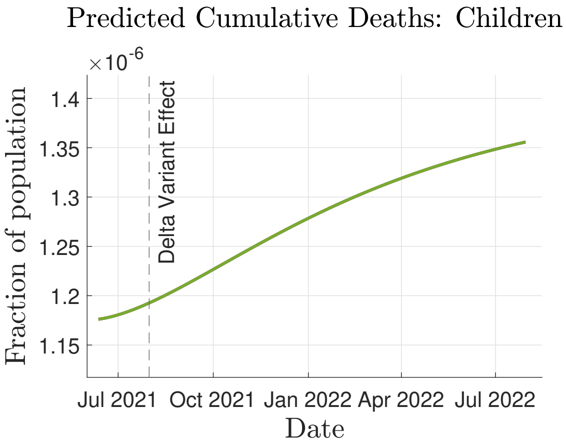


(a) Children

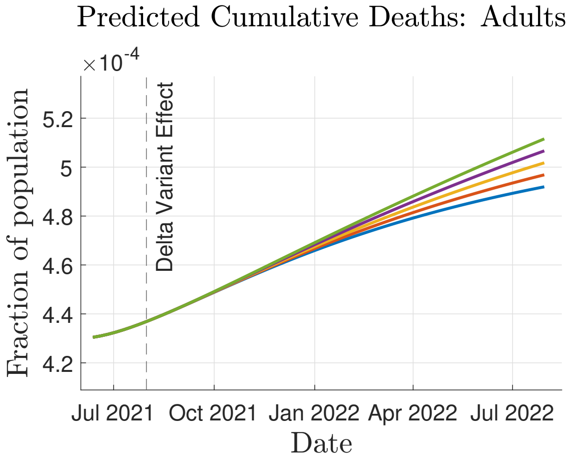


(b) Adults

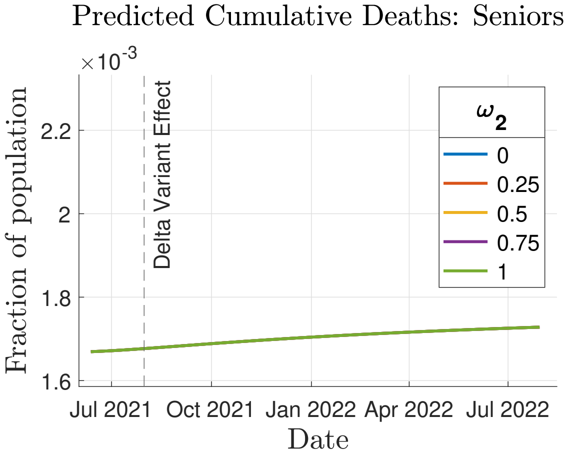


(c) Seniors

FIG. S19: Simulation Case 3: Effect of Anti/Non-Vaxxers on COVID-19 Transmission. Variation of adult Anti/Non-Vaxxer proportion, $\omega_2$, with $\omega_1 = 0$ (children) and $\omega_3 = 0$ (seniors). Vaccine inefficacy, $\sigma = 0.05$, and transmissibility parameter, $K_{new}/K = 1$. Simulated Deaths for a) children, b) adults, and c) seniors.

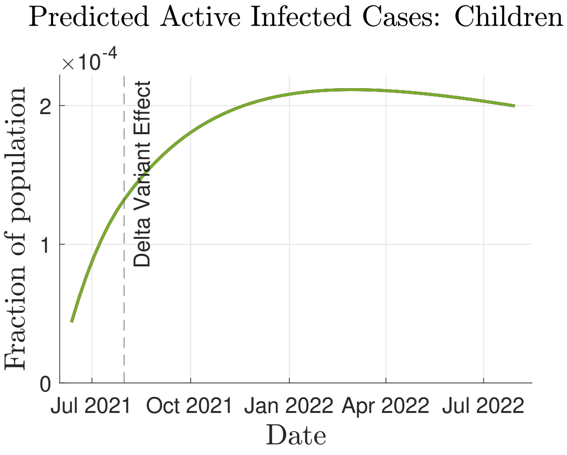


(a) Children

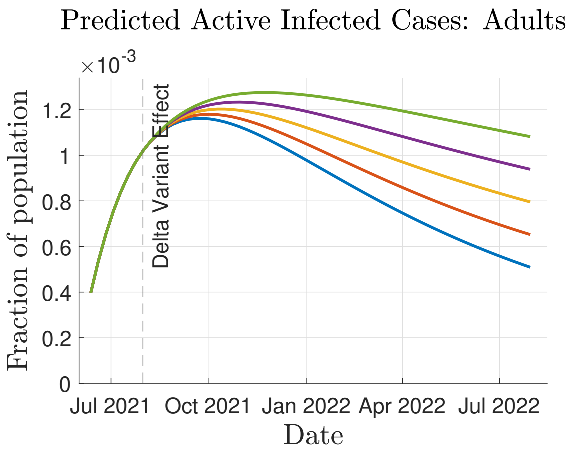


(b) Adults

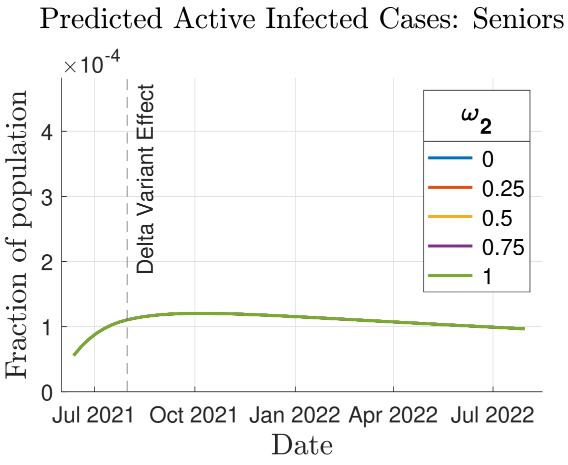


(c) Seniors

FIG. S20: Simulation Case 3: Effect of Anti/Non-Vaxxers on COVID-19 Transmission. Variation of adult Anti/Non-Vaxxer proportion, $\omega_2$, with $\omega_1 = 1$ (children) and $\omega_3 = 1$ (seniors). Vaccine inefficacy, $\sigma = 0.05$, and transmissibility parameter, $K_{new}/K = 1$. Simulated Infections for a) children, b) adults, and c) seniors.

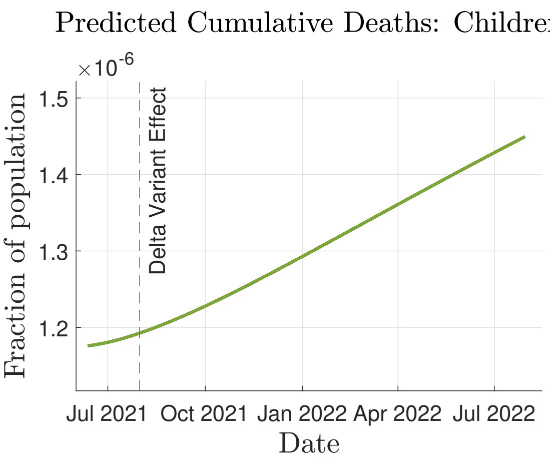


(a) Children

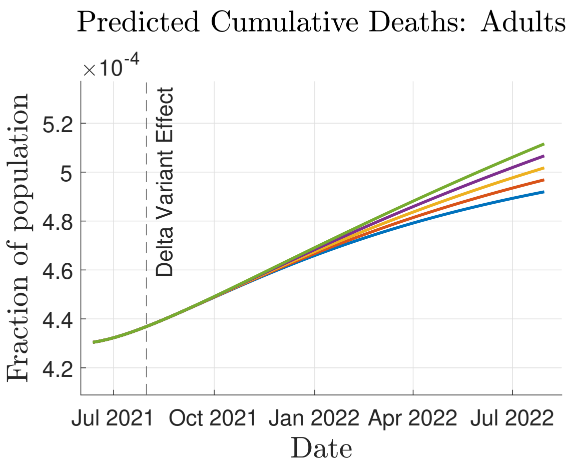


(b) Adults

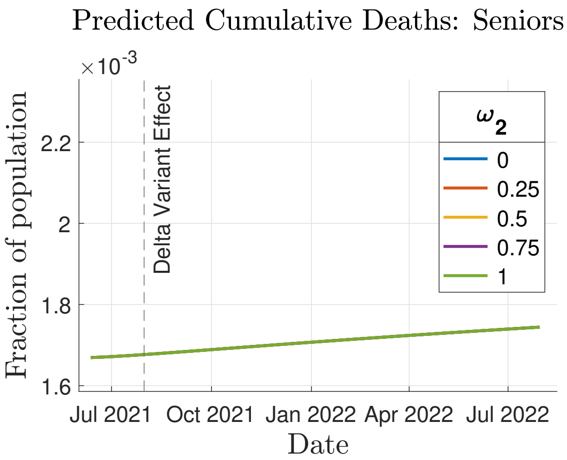


(c) Seniors

FIG. S21: Simulation Case 3: Effect of Anti/Non-Vaxxers on COVID-19 Transmission. Variation of adult Anti/Non-Vaxxer proportion, $\omega_2$, with $\omega_1 = 1$ (children) and $\omega_3 = 1$ (seniors). Vaccine inefficacy, $\sigma = 0.05$, and transmissibility parameter, $K_{new}/K = 1$. Simulated Deaths for a) children, b) adults, and c) seniors.

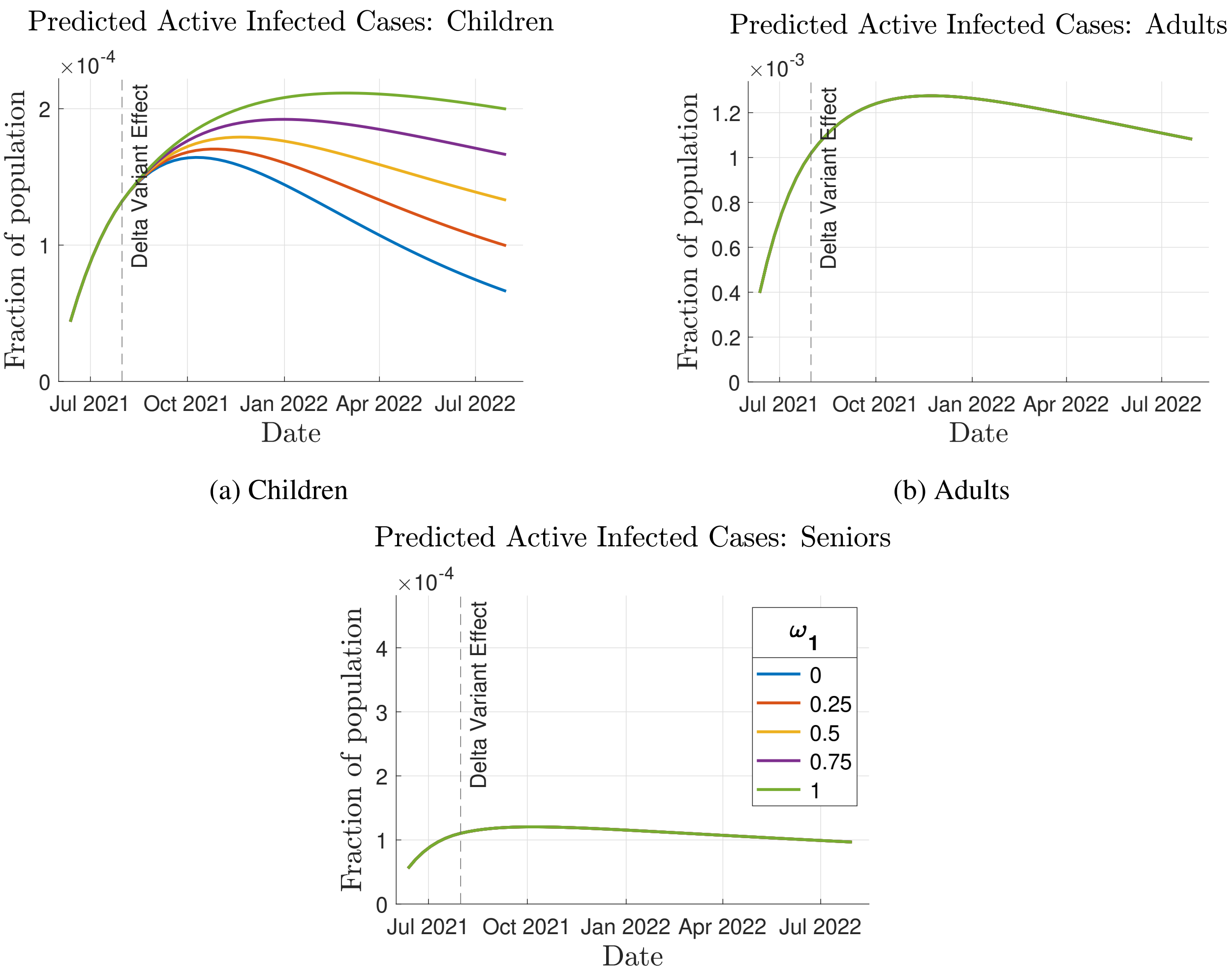


(a) Children

(b) Adults

(c) Seniors

FIG. S22: Simulation Case 3: Effect of Anti/Non-Vaxxers on COVID-19 Transmission. Variation of children Anti/Non-Vaxxer proportion, $\omega_1$, with $\omega_2 = 1$ (adults) and $\omega_3 = 1$ (seniors). Vaccine inefficacy, $\sigma = 0.05$, and transmissibility parameter, $K_{new}/K = 1$. Simulated Infections for a) children, b) adults, and c) seniors.

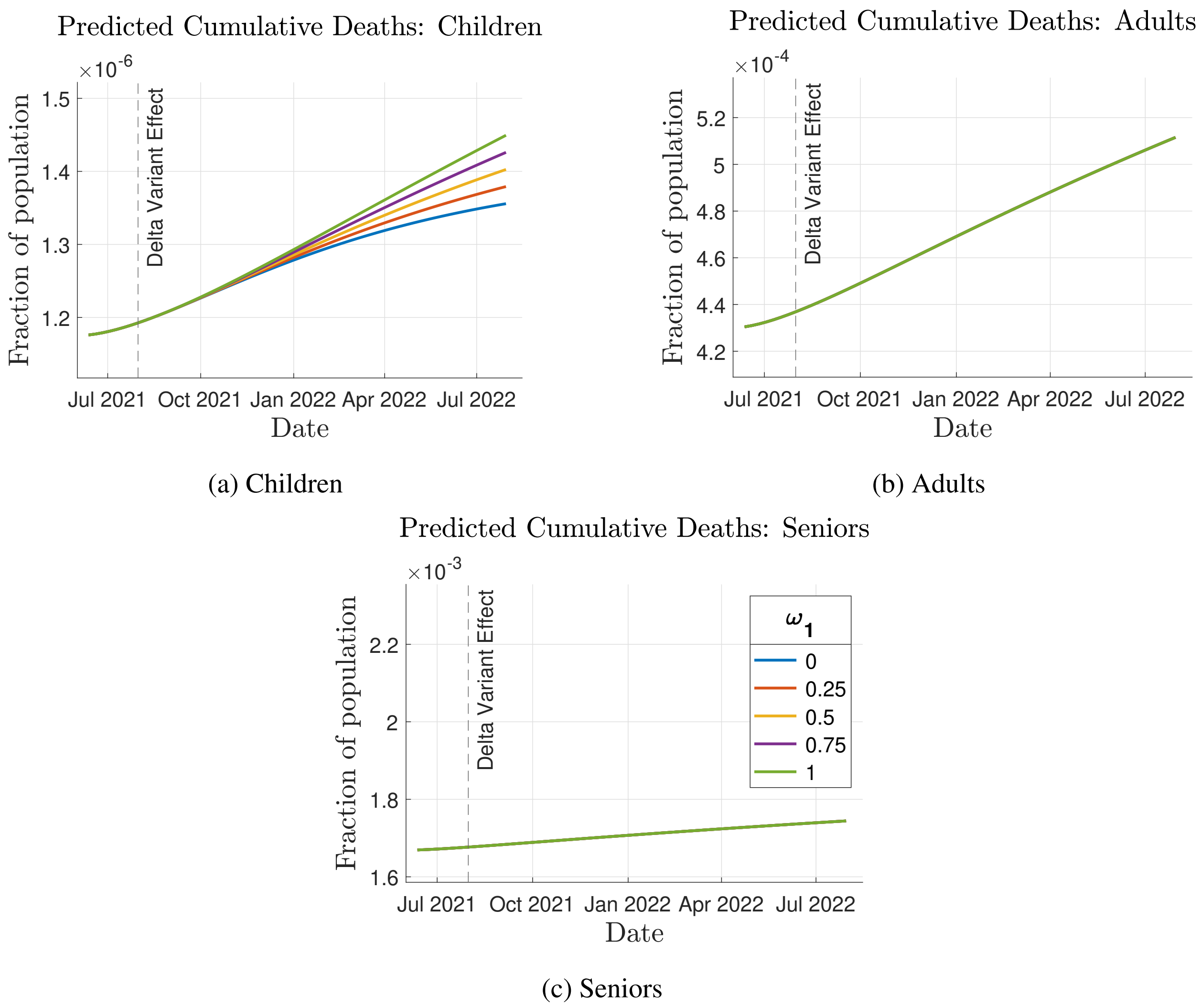


FIG. S23: Simulation Case 3: Effect of Anti/Non-Vaxxers on COVID-19 Transmission. Variation of children Anti/Non-Vaxxer proportion, $\omega_1$, with $\omega_2 = 1$ (adults) and $\omega_3 = 1$ (seniors). Vaccine inefficacy, $\sigma = 0.05$, and transmissibility parameter, $K_{new}/K = 1$. Simulated Deaths for a) children, b) adults, and c) seniors.

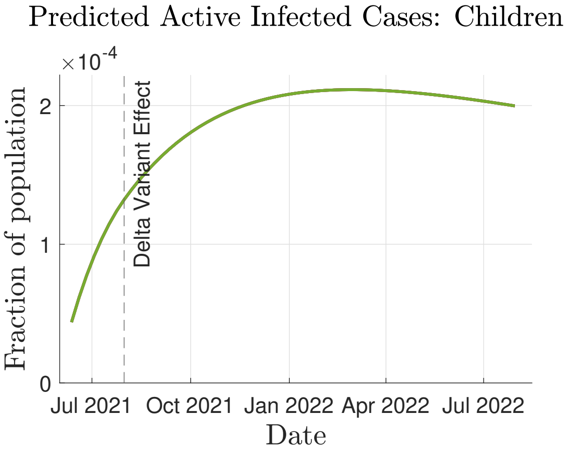


(a) Children

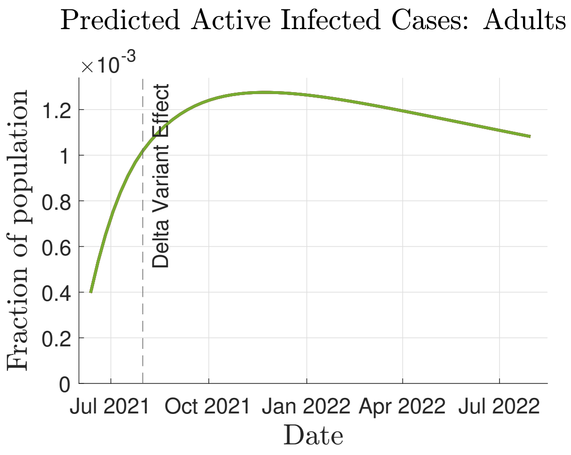


(b) Adults

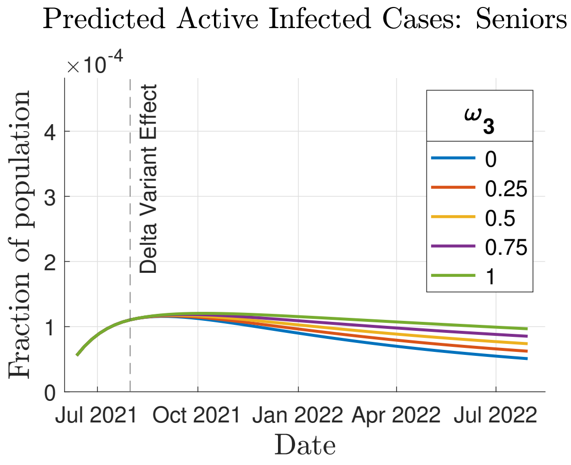


(c) Seniors

FIG. S24: Simulation Case 3: Effect of Anti/Non-Vaxxers on COVID-19 Transmission. Variation of senior Anti/Non-Vaxxer proportion, $\omega_3$, with $\omega_1 = 1$ (children) and $\omega_2 = 1$ (adults). Vaccine inefficacy, $\sigma = 0.05$, and transmissibility parameter, $K_{new}/K = 1$. Simulated Infections for a) children, b) adults, and c) seniors.

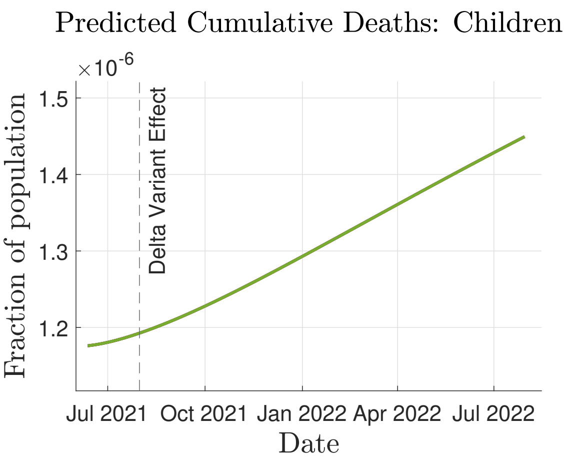


(a) Children

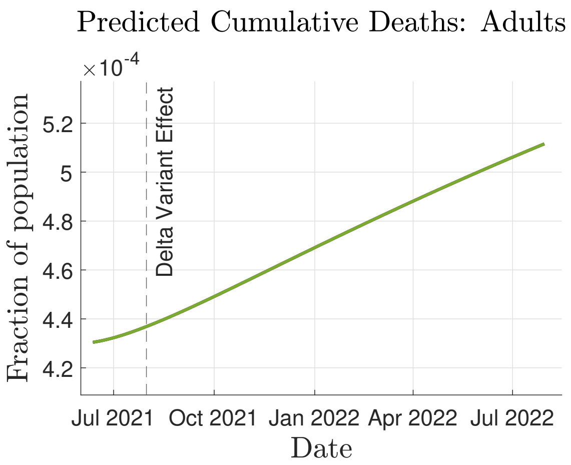


(b) Adults

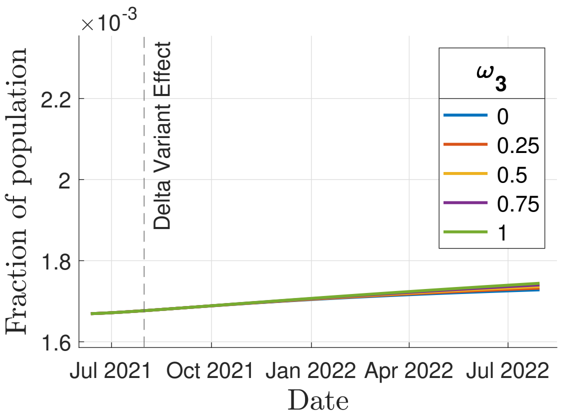


(c) Seniors

FIG. S25: Simulation Case 3: Effect of Anti/Non-Vaxxers on COVID-19 Transmission. Variation of senior Anti/Non-Vaxxer proportion, $\omega_3$, with $\omega_1 = 1$ (children) and $\omega_2 = 1$ (adults). Vaccine inefficacy, $\sigma = 0.05$, and transmissibility parameter, $K_{new}/K = 1$. Simulated Deaths for a) children, b) adults, and c) seniors.
